# SARS-CoV-2 anti-spike IgG antibody responses after second dose of ChAdOx1 or BNT162b2 and correlates of protection in the UK general population

**DOI:** 10.1101/2021.09.13.21263487

**Authors:** Jia Wei, Koen B. Pouwels, Nicole Stoesser, Philippa C. Matthews, Ian Diamond, Ruth Studley, Emma Rourke, Duncan Cook, John I Bell, John N Newton, Jeremy Farrar, Alison Howarth, Brian D. Marsden, Sarah Hoosdally, E Yvonne Jones, David I Stuart, Derrick W. Crook, Tim E. A. Peto, A. Sarah Walker, David W. Eyre, COVID-19 Infection Survey team

**Author notes:** contribution considered equal (Jia Wei, Koen B. Pouwels). contribution considered equal (A. Sarah Walker, David W. Eyre). See Acknowledgements for the Coronavirus Infection Survey team. Corresponding author: Dr David Eyre, Microbiology Department, John Radcliffe Hospital, Headley Way, Oxford, OX3 9DU.

## Abstract

We investigated anti-spike IgG antibody responses and correlates of protection following second doses of ChAdOx1 or BNT162b2 SARS-CoV-2 vaccines in the UK general population. In 222,493 individuals, we found significant boosting of anti-spike IgG by second doses of both vaccines in all ages and using different dosing intervals, including the 3-week interval for BNT162b2. After second vaccination, BNT162b2 generated higher peak levels than ChAdOX1. Older individuals and males had lower peak levels with BNT162b2 but not ChAdOx1, while declines were similar across ages and sexes with ChAdOX1 or BNT162b2. Prior infection significantly increased antibody peak level and half-life with both vaccines. Anti-spike IgG levels were associated with protection from infection after vaccination and, to an even greater degree, after prior infection. At least 67% protection against infection was estimated to last for 2-3 months after two ChAdOx1 doses and 5-8 months after two BNT162b2 doses in those without prior infection, and 1-2 years for those unvaccinated after natural infection. A third booster dose may be needed, prioritised to ChAdOx1 recipients and those more clinically vulnerable.

## Introduction

The Pfizer-BioNTech BNT162b2 and Oxford-AstraZeneca ChAdOx1 nCoV-19 (hereafter ChAdOx1) SARS-CoV-2 vaccines have been widely used in the United Kingdom and worldwide^1,2^. In the UK, vaccines were initially prioritised to older adults, frontline healthcare and social-care workers, and clinically vulnerable individuals, and then offered to other adults in decreasing age order^3^. To 4^th^ October 2021, 85% and 78% of the population (aged ≥12y) have received one and two doses, respectively^4^.

With wide-spread Alpha variant transmission, in January 2021 the UK government extended the dosing interval from 3-4 to 12 weeks for all vaccines to maximize first dose coverage, based on preliminary data showing high short-term efficacy from single BNT162b2 (90%) and ChAdOx1 (70%) doses^5^. This approach raises several questions. Although the ChOxAd1 trial found higher vaccine efficacy with dosing intervals ≥6 weeks^6^, BNT162b2 trials did not compare different dosing intervals. Subsequent UK studies showed extended BNT162b2 dosing intervals generated a higher antibody response than the 3-week interval^7–9^. However, these studies were based on relatively small sample sizes (N<600) or specific population groups such as healthcare workers, potentially reducing generalisability, and antibody levels were only measured at specific times after second doses. With the rapid emergence of the Delta variant, and greater protection after second than first vaccine doses^10–12^, from mid-2021 dosing intervals were reduced to 8 weeks^13^ to achieve greater protection faster. However, large population-based investigations of how these different dosing intervals, or other factors, affect longer-term antibody changes after the second dose are limited, but essential to assess the duration of protection and the need for booster doses.

Following the ChAdOx1 vaccine, anti-spike IgG and pseudovirus neutralisation titres are associated with protection against symptomatic SARS-CoV-2 infection^14^. Similarly for the mRNA-1273 vaccine, anti-spike antibody and neutralisation titres were inversely associated with SARS-CoV-2 infection, with 68.5% of vaccine efficacy up to 126 days post-second dose mediated by day 29 neutralisation titres^15^. However, how these measurements relate to antibody levels and durations of protection in populations over time, and following BNT162b vaccination, is not fully understood.

We used data from the UK’s national COVID-19 Infection Survey (ISRCTN21086382), a large representative sample of households with longitudinal follow-up, to investigate longer-term anti-trimeric spike IgG antibody responses following second ChAdOx1 or BNT162b2 vaccinations, and quantified the impact of dosing interval, age, and prior infection status on antibody peak levels and declines. We estimated the association between anti-spike IgG levels and protection against SARS-CoV-2 infection, and combined these findings with estimated antibody trajectories to predict the duration of protection following second vaccination and natural infection.

## Results

From 8^th^ December 2020 to 4^th^ October 2021, 222,493 participants received two ChAdOx1 or two BNT162b2 vaccinations and had at least one antibody measurement from 91 days before the first vaccination onwards. The median (interquartile range, IQR) age was 57 (43-68) years, 120,866 (54.3%) were female, and 209,898 (94.3%) reported white ethnicity. 7,071 (3.2%) reported working in patient-facing healthcare, and 62,814 (28.2%) having a long-term health condition. 121,322 (54.5%) and 79,693 (35.8%) participants without evidence of prior infection (see Methods) received two doses of ChAdOx1 or BNT162b2, as did 12,066 (5.4%) and 9,412 (4.2%) with evidence of prior infection, respectively. These four cohorts contributed 723,844 anti-spike IgG measurements **(Extended Data Fig. 1)**. The median (IQR) [range] dosing interval was 76 (68-78) [17-237] and 76 (66-78) [17-225] days for those receiving ChAdOx1 without or with prior infection, respectively, and 71 (58-77) [17-289] and 65 (56-76) [17-238] days respectively for BNT162b2 **(Supplementary Table 1)**.

### Anti-spike IgG response following first and second dose

In participants receiving two vaccinations without prior infection, generalised additive models (GAMs) adjusting only for age and dosing interval showed generally similar antibody trajectories for both vaccines, but with higher antibody levels achieved with BNT162b2 vs. ChAdOx1 **(Extended Data Fig. 2a, c, Supplementary Table 2a; observed data: Supplementary Fig. 1)**. Anti-spike IgG levels increased after the first dose, peaked ∼21 days later, then gradually declined until the second dose, after which they reached even higher peak levels ∼21 days later, then gradually declined again. Post-first dose, peak levels were lower in older participants, but age differences were attenuated after the second dose **(Extended Data Fig. 3a**,**c, Supplementary Table 2b)**. In these minimally adjusted analyses (adjusted for age and dosing interval), there was no evidence of differences in antibody levels and declines after the second dose across 8-week to 12-week dosing intervals for both ChAdOx1 and BNT162b2 vaccines. However, the antibody trajectories were different for the 3-week BNT162b2 dosing interval, where antibody levels gradually increased from the start of the first dose until around 42 days after the second dose, after which antibody levels were generally similar to those with 8–12-week dosing intervals, although slightly lower in 80-year-olds **(Extended Data Fig. 2c)**.

In participants with evidence of prior infection, antibody levels started from levels above 23 BAU/mL (the positivity threshold, see Methods) and gradually increased for both vaccines; there was no evidence of differences in antibody levels and declines after the second dose for dosing intervals from 8 to 12 weeks **(Extended Data Fig. 2b, d; observed data shown in Supplementary Fig. 2)**. Following prior infection, antibody levels rose to lower levels in older vs. younger participants after the first dose, but the difference was attenuated after the second dose **(Extended Data Fig. 3b, d)**. In those with prior infection, there was less boosting seen from the second dose given higher levels after one dose, compared to those not previously infected, but there was a second dose boosting effect for 80-year-olds **(Fig. 1)**. For ChAdOx1, participants without prior infection had lower antibody levels post-second dose than those with prior infection post-first dose; but for BNT162b2, two vaccinations without prior infection led to higher antibody levels than previously infected participants having only one dose, especially for 80-year-olds **(Fig. 1**; trajectories for other dosing intervals in **Extended Data Fig. 4)**.

**Fig. 1.**
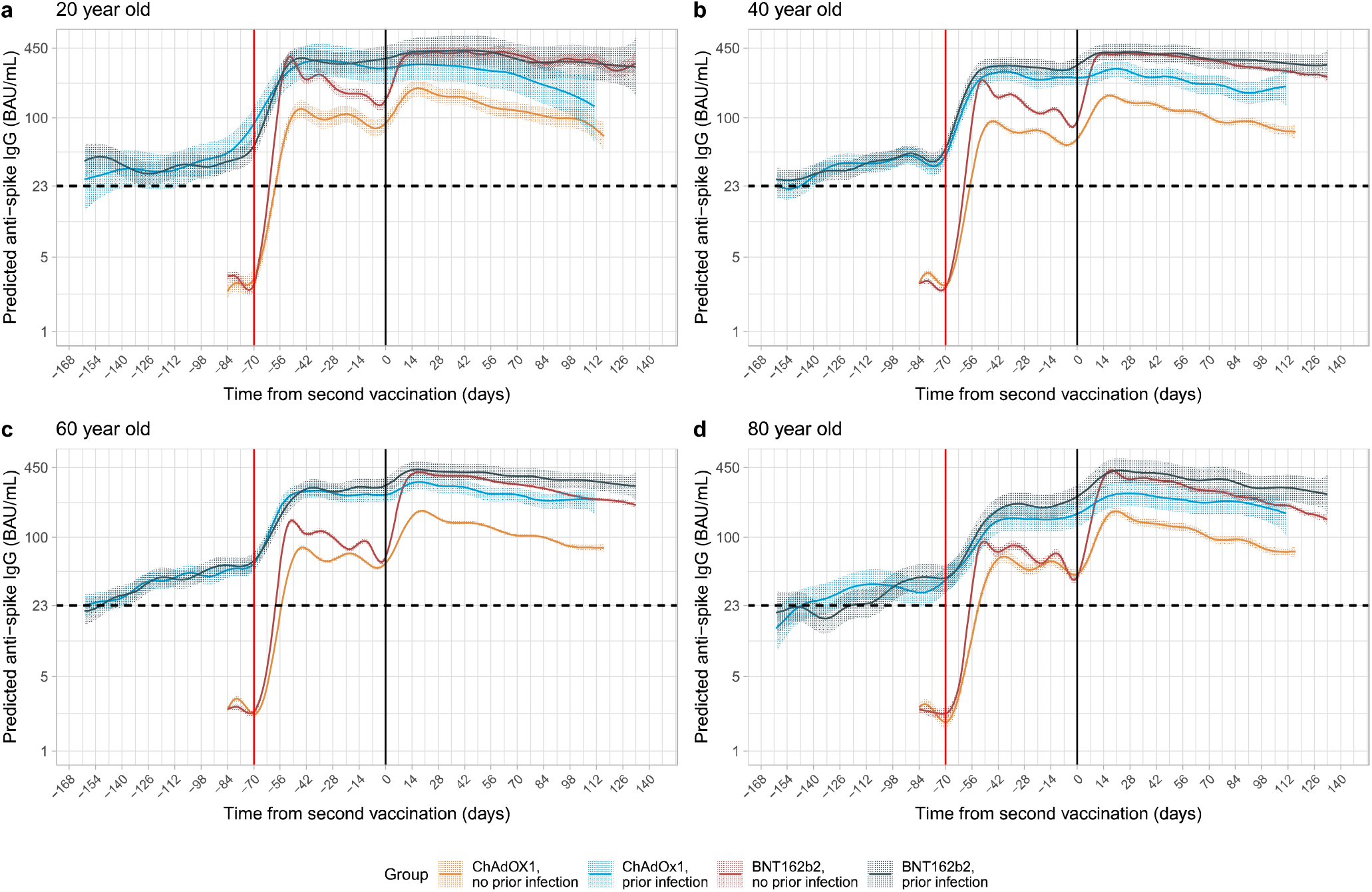
Predicted anti-spike IgG levels in participants with 10-week dosing interval by time from second vaccination according to vaccine type, age and prior infection status using generalised additive models adjusted for age and dosing interval. **a**, 20-year-old. **b**, 40-year-old. **c**, 60-year-old. **d**, 80-year-old. Predicted levels are plotted on a log scale. Black dotted line indicates the threshold of IgG positivity (23 BAU/mL). Red solid line indicates the first vaccination and black solid line indicates the second vaccination. Line colour indicates response predicted for ChAdOx1 and BNT162b2, with or without prior infection. See **Extended Data Fig. 3** for a replotted version of the same estimates to allow comparison by age for each vaccine type and prior infection status, see **Extended Data Fig. 4** for 8 and 12 week dosing interval. The 95% CIs are calculated by prediction ± 1.96 × standard error of the prediction.

### Determinants of anti-spike IgG peak levels and half-life after the second dose

#### ChAdOx1

Of the 133,388 participants (with or without prior infection) who received two ChAdOx1 doses, 100,639 participants contributed 191,137 antibody measurements ≥21 days after the second dose, median (IQR) [range] 2 (1-2) [1-5] measurements per participant (**Supplementary Table 3)**. Antibody measurements were taken a median (IQR) [range] 61 (41-83) [21-119] days post second vaccination (**Supplementary Table 3, Supplementary Fig. 3**). Assuming antibody levels declined exponentially, using Bayesian linear mixed models we estimated a mean peak anti-spike IgG level of 184 BAU/mL (95% credible interval, Crl 183-185), and mean half-life of 79 days (78-80) **(Extended Data Fig. 5)**, versus 113 BAU/mL (106-117) and 184 days (163-210) respectively following natural infection before vaccination^16^. There was no evidence that rates of antibody decline flattened over time (up to 119 days post second vaccination, see Methods). In a multivariable model, all factors considered (age, sex, ethnicity, reporting a long-term health condition, healthcare work, deprivation, dosing interval and prior infection status) were independently associated with anti-spike IgG peak levels 21 days post-second dose, but most effects were small **(Fig. 2, Supplementary Table 4)**. The largest effects were associated with prior infection, peak 219 BAU/mL higher (210-227) and ethnicity, peak 41 BAU/mL (36-47) higher in those reporting non-white ethnicity. Peak levels were slightly lower in males, those reporting a long-term health condition, those not working in healthcare, with shorter dosing intervals, younger age, and less deprivation (higher deprivation percentile). Prior infection extended the half-life by 13 days (95%Crl 9-17). There were very small reductions in half-life at older ages, with non-white ethnicity, and having a long-term health condition. There was no evidence of associations between half-life and sex, being a healthcare worker, and dosing interval in participants who received ChAdOx1.

**Fig. 2.**
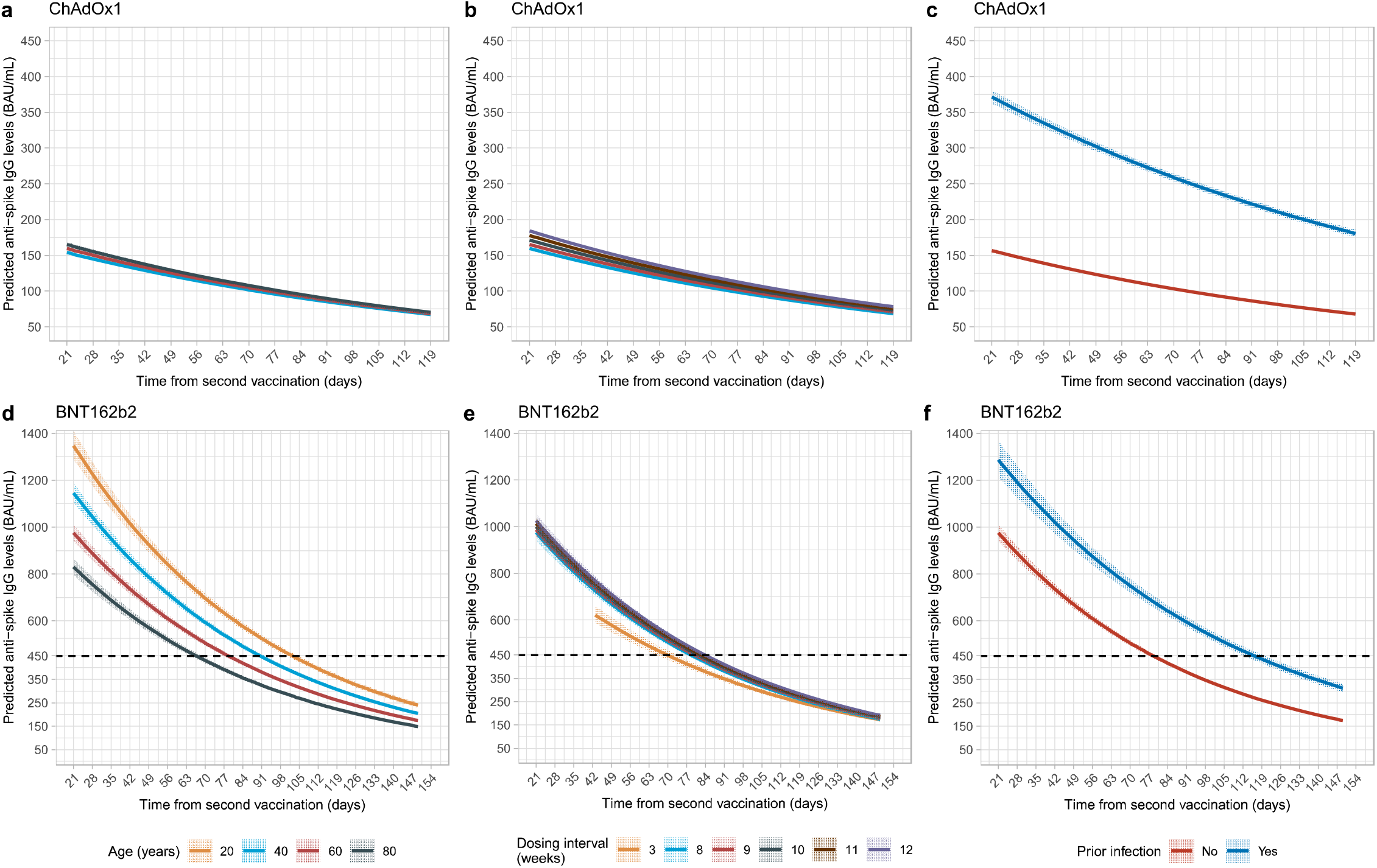
Posterior predicted trajectories of anti-spike IgG levels from 21 days post-second dose by age (panel a, d), dosing interval (panel b, e), and prior infection status (panel c, f) using Bayesian linear mixed interval censored models adjusted for age, sex, ethnicity, long-term health conditions, healthcare role, deprivation, dosing interval, and prior infection status. **Panel a**,**b**,**c** show participants who received two doses of ChAdOx1. **Panel d**,**e**,**f** show participants who received two doses of BNT162b2. Black dotted line shows the upper quantification limit of 450 BAU/mL. For BNT162b2, as 52% measurements were above the upper limit, the estimated peak levels were higher than 450 BAU/mL (interval censoring accounted for in analysis). Plotted at reference categories: 60 years, female, white ethnicity, not reporting a long-term health condition, not a healthcare worker, deprivation percentile=60, 8-week dosing interval, and no prior infection. In **panel a**, 20-year-old group is not plotted because the vast majority of those receiving ChAdOx1 were ≥40 years. These model estimates more completely adjust for confounders than the generalised additive models plotted in Extended Data Fig. 2, but only estimate trends after second vaccination.

#### BNT162b2

In 89,105 participants (with or without prior infection) who received two BNT162b2 doses, 55,053 participants contributed 120,728 antibody measurements ≥21 days after the second dose (≥42 days for those with 3-week dosing interval, see Methods, **Extended Data Fig. 2**), median (IQR) [range] 2 (1-3) [1-6] per participant (**Supplementary Table 3**). Antibody measurements were taken a median (IQR) [range] 79 (51-106) [21-149] days post second vaccination (**Supplementary Table 3, Supplementary Fig. 3**). The estimated mean peak level was 959 BAU/mL (95%Crl 944-974), and the mean half-life was 51 days (50-52) **(Extended Data Fig. 5)**. There was again no evidence of antibody decline flattening on the log scale (up to 149 days post second vaccination, see Methods). Factors had greater effects on peak levels for BNT162b2 than ChAdOx1 **(Fig. 2, Supplementary Table 4)**. Peak levels were lower at older ages (76 BAU/mL lower per 10-years older, 95%Crl 68-84), in males (140 BAU/mL lower, 117-164), those reporting long-term health conditions (79 BAU/mL lower, 55-104), and were higher in those reporting non-white ethnicity (141 BAU/mL higher, 78-208), working in healthcare (287 BAU/mL higher, 221-358), and less deprived (5 BAU/mL higher per 10 percentiles higher, 1-10). However, these factors had little or no effect on half-life. Within dosing intervals between 8 and 12 weeks, longer dosing intervals were associated with higher peak levels (12 BAU/mL higher per week longer, 1-23), but had no effect on half-life. Compared with an 8-week extended schedule, a 3-week dosing interval was associated with a lower peak level 42 days post-second dose but a slightly longer half-life (6 days longer, 2-10), leading to similar antibody levels at 149 days across different dosing groups. Similar to ChAdOX1, prior infection was associated with a much higher peak level (312 BAU/mL higher, 248-381) and a longer half-life (11 days longer, 8-15).

Comparing the effects of factors between the two vaccines, and with our previous findings on natural infection^16^ **(Fig. 3)**, effects of some factors were relatively consistent between ChAdOx1, BNT162b2 and/or natural infection, albeit with differing effect sizes (e.g sex, ethnicity, long-term health condition, working in healthcare, dosing interval on peak; prior infection on peak and half-life), whilst for others effects were in opposite directions (e.g age, deprivation on peak) or only associated for one vaccine (e.g. ethnicity, working in healthcare on half-life). In general, effects on peak levels were greater for BNT162b2 than ChAdOx1; and, other than prior infection, effects on half-life were limited for both vaccines.

**Fig. 3.**
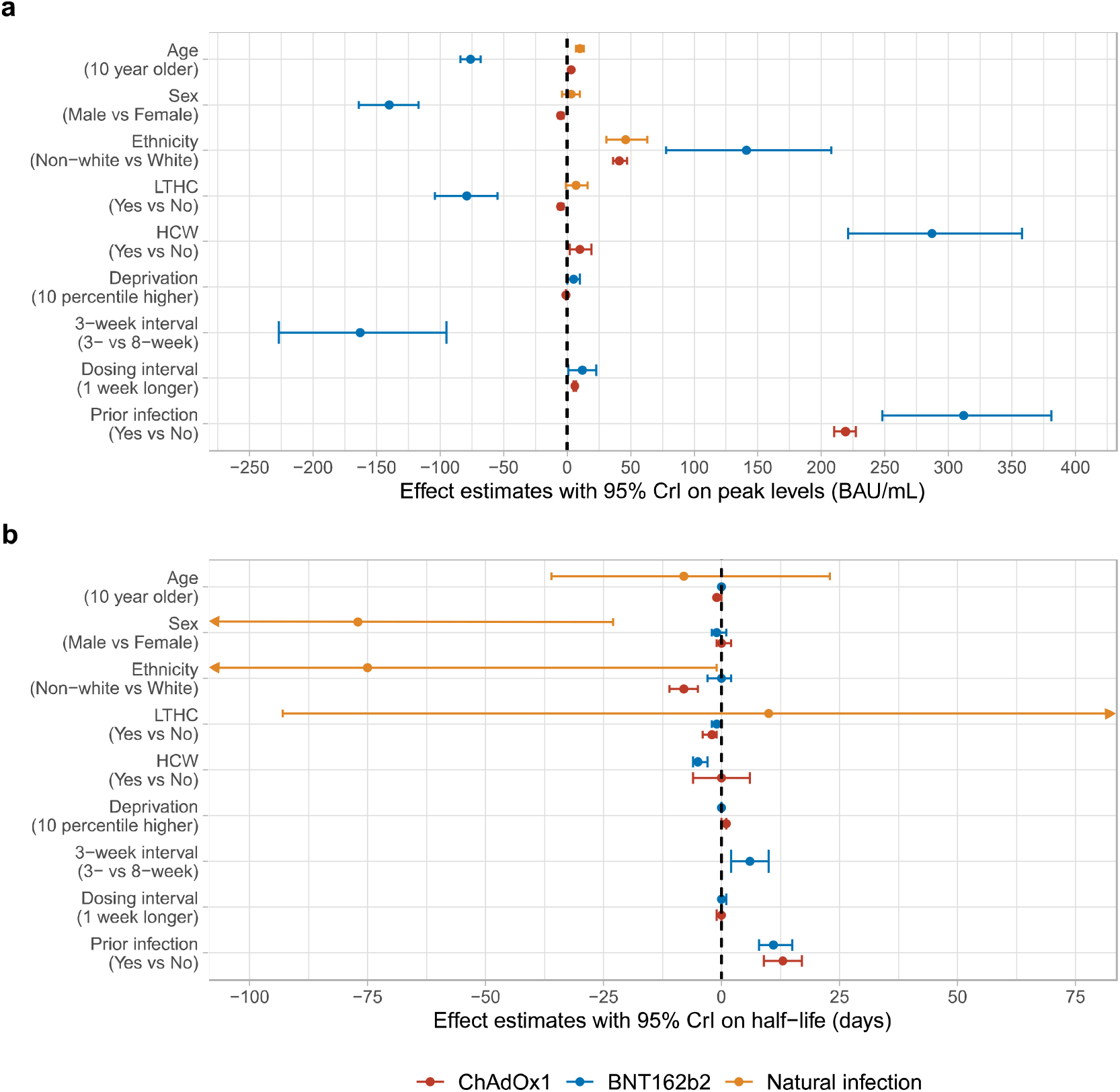
Comparison of effect of factors in participants who received two doses of ChAdOx1 or BNT162b2 or had natural SARS-CoV-2 infection. **a**, Effect on anti-spike IgG peak levels. **b**, Effect on anti-spike IgG half-lives. Mean estimates with 95% credible intervals are presented. In panel b, 95% credible intervals are truncated at -100 and 75 days for visualisation.

### Correlates of protection

To interpret anti-spike IgG levels over time, we investigated whether recent antibody levels are correlates of protection from infection following prior infection or vaccination in the general population. We used data from 17^th^ May 2021 to 4^th^ October 2021, i.e. while the Delta variant accounted for nearly all cases^17^, and fitted logistic generalised additive models for detected new infections, investigating the effect of the most recent antibody measurement obtained 21-59 days earlier on PCR test results at each study visit (distribution of visits relative to first vaccination shown in **Supplementary Fig. 4** and **Supplementary Table 5**). Three groups were investigated, unvaccinated participants with or without evidence of prior infection (6833 participants; 12,560 visits), participants vaccinated with ChAdOx1 ≥21 days previously (83,924 participants; 221,380 visits, 36-273 days post-first vaccination), and participants vaccinated with BNT162b2 ≥21 days previously (49,820 participants; 124,822 visits, 35-298 days post-first vaccination). Participants tested PCR-positive at 202 (1.6%), 1327 (0.6%), and 591 (0.5%) visits, respectively. Vaccinated participants with evidence of prior infection were excluded, as there were insufficient data to model these groups separately, and associations may differ. Adjustment was made for confounders, including age, geography, and calendar time (see Methods).

Compared with unvaccinated individuals with a most recent anti-spike IgG measurement of 1 BAU/mL, protection against infection increased steeply as antibody levels rose in unvaccinated participants, consistent with protection from prior infection, with 50% protection at 20 BAU/mL (below the 23 BAU/mL positivity threshold) and 67% protection at 33 BAU/mL. Higher antibody levels were needed to achieve the same level of protection post vaccination, with no evidence of differences in protection between ChAdOx1 and BNT162b2 at any given antibody level (**Fig. 4a**). For example, considering antibody levels where two thirds (67%) of individuals were protected against infection, participants vaccinated with ChAdOx1 or BNT162b2 required estimated levels of 107 BAU/mL and 94 BAU/mL (100 BAU/mL in models pooling both vaccines), respectively. However, as described above, antibody levels rose to higher levels following BNT162b2 than ChAdOx1, explaining higher vaccine effectiveness after BNT162b vs. ChAdOx1^18^, with natural infection resulting in the lowest antibody levels but the greatest protection at a given antibody level (**Fig. 4d**,**e**,**f**). Protection against infection with moderate to high viral loads (Ct values <30) (**Fig. 4b**) and symptomatic infection (**Fig. 4c**) was similar. Findings were also similar when considering the maximum prior antibody measurement (which had a worse model fit), in part because of the limited time for antibody waning to occur between maximum and most recent antibody measurements (**Extended Data Fig. 6**).

**Fig. 4.**
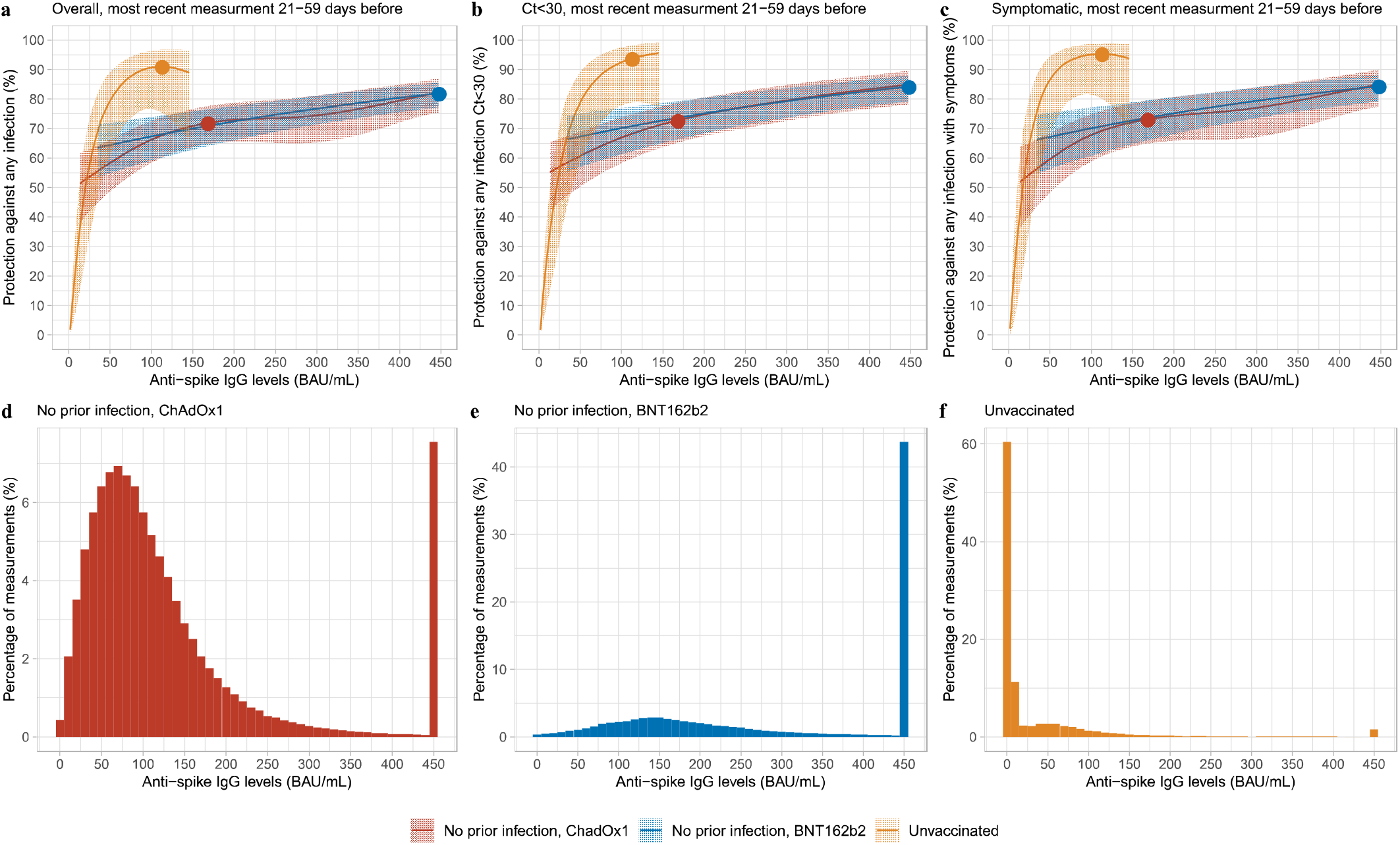
Association between anti-spike IgG levels and protection from SARS-CoV-2 infection using the most recent antibody measurement obtained 21-59 days prior to the current visit. **a**, protection against any infection; **b**, protection against infection with a moderate to high viral load (Ct value <30); **c**, protection against infection with self-reported symptoms. Three groups are investigated, unvaccinated participants with or without evidence of prior infection, participants vaccinated with ChAdOx1 without evidence of prior infection, and participants vaccinated with BNT162b2 without evidence of prior infection. Dots represent the median predicted individual peak levels from the Bayesian linear mixed models: 1026 BAU/mL for BNT162b2 (plotted at the upper quantification limit of 450 BAU/mL), 167 BAU/mL for ChAdOx1, and 111 BAU/mL for unvaccinated participants^16^. Distribution of the most recent anti-spike IgG measurements for the three population groups are shown in panel **d, e, f**. See Supplementary Fig. 4 and Supplementary Table 5 for timing of visits relative to first vaccination.

### Duration of antibody response and association with protection

Models of correlates of protection allowed post-vaccination antibody levels corresponding to 67% protection from Delta variant infection to be estimated (data were insufficient to estimate lower protection percentages for BNT162b). The estimated mean time from second vaccination to antibody measurements reaching levels associated with 67% protection was 55-86 days for ChAdOx1 vaccinated participants without prior infection, with limited variation across age, sex, dosing interval, or long-term health conditions (**Fig. 5a**). For BNT162b2, the estimated mean durations were 161-227 days, and were longer at younger ages, in females, those without long-term health conditions, but were similar across different dosing intervals (**Fig. 5b**). For unvaccinated participants infected previously, using a model of antibody declines after natural infection^16^, the duration from diagnosis to the level associated with 67% protection was estimated to be 1-2 years (**Fig. 5c**). Older people had a longer duration of protection after natural infection, but these results were conditional on seroconverting, and older individuals had lower seroconversion rates (**Fig. 5d**). Although data were insufficient to estimate antibody levels correlated with protection for vaccinated participants with prior infection, conservatively assuming threshold levels were similar to those vaccinated without prior infection and given their half-lives were longer, the duration of protection could last for >1 year. Times for antibody levels to fall to the threshold for positivity, i.e. 23 BAU/mL, were longer, but followed the same patterns (**Extended Data Fig. 7**).

**Fig. 5.**
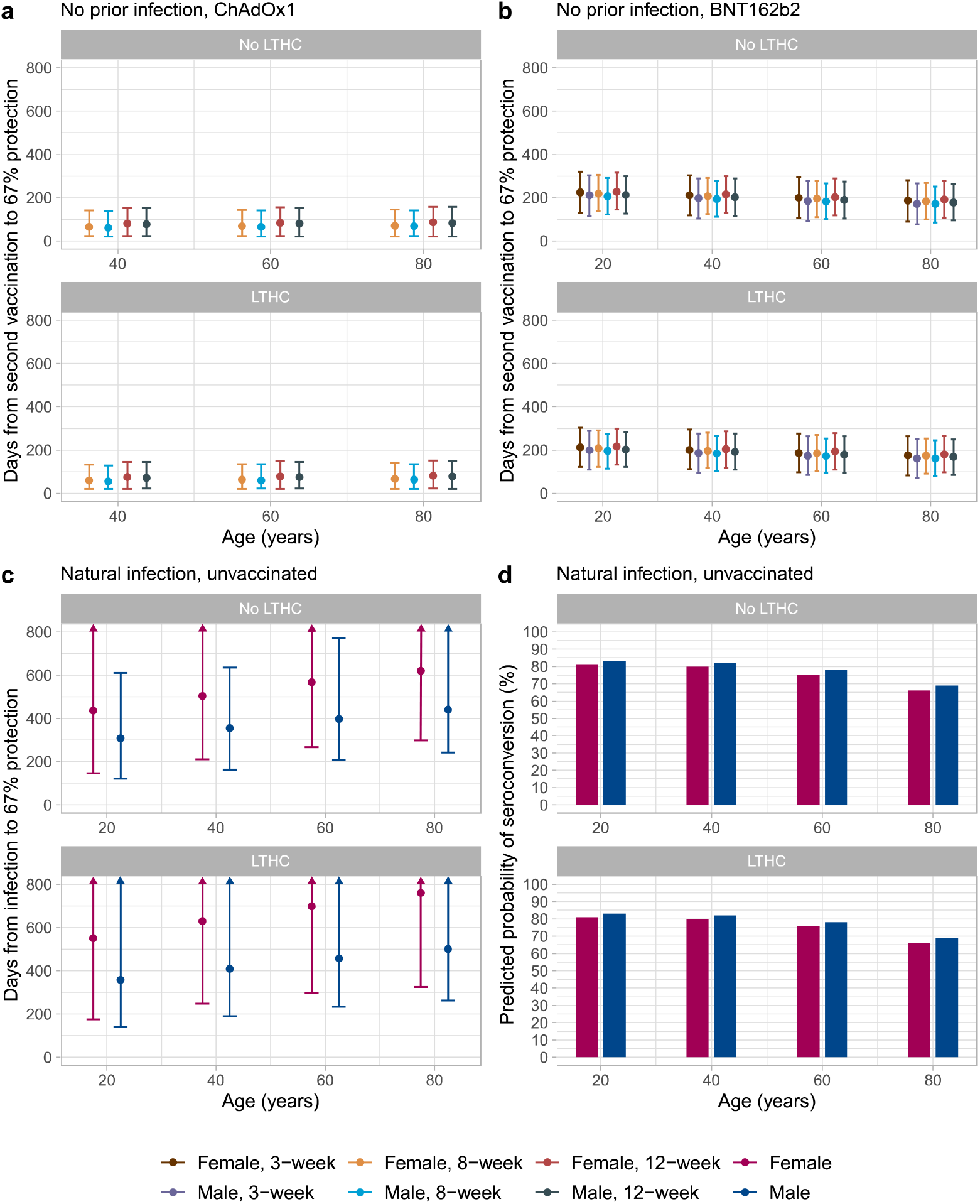
Posterior predicted days (95% credible interval) from the second vaccination/infection to the threshold level associated with 67% protection (ChAdOx1: 107 BAU/mL, BNT162b2: 94 BAU/mL, unvaccinated: 33 BAU/mL). **a**, In participants without prior infection and vaccinated with ChAdOX1. **b**, In participants without prior infection and vaccinated with BNT162b2. **c**, In unvaccinated participants who had natural infection. **d**, Predicted probability of seroconverting in unvaccinated participants who had natural infection, based on a previous model on seroconversion^16^. Estimates were separated by age, sex, dosing interval, long-term health condition (LTHC), and vaccine type for vaccinated people, and by age, sex, and LTHC for unvaccinated people. y-axis is truncated at 800 days for visualisation. For ChAdOx1, 20-year-old group is not plotted because the vast majority of those receiving ChAdOx1 were ≥40 years. Panel **a**,**b**,**c** are conditional on participants having antibody response/seroconverting (see section of “Vaccine non-responders” for ChAdOx1 and BNT162b2, and discussion in a previous publication^16^ for unvaccinated individuals).

We estimated the proportion of participants with 67% protection at 90, 180, 270, and 360 days from second dose or natural infection based on individual-level predictions, conditional on seroconverting. At 180 days, 10% of ChAdOx1 participants without prior infection remained above the level associated with 67% protection, with little variation across different factors. For BNT162b2, the proportion varied between 40-80% for those without prior infection and was higher in younger ages, females, and those without long-term health conditions. Over 90% of unvaccinated participants with natural infection were above the level required for 67% protection 180 days post diagnosis. At 270 days, almost everyone receiving ChAdOx1 or BNT162b2 fell below the level of 67% protection, while over 80% of unvaccinated participants were still above this threshold level (**Fig. 6**).

**Fig. 6.**
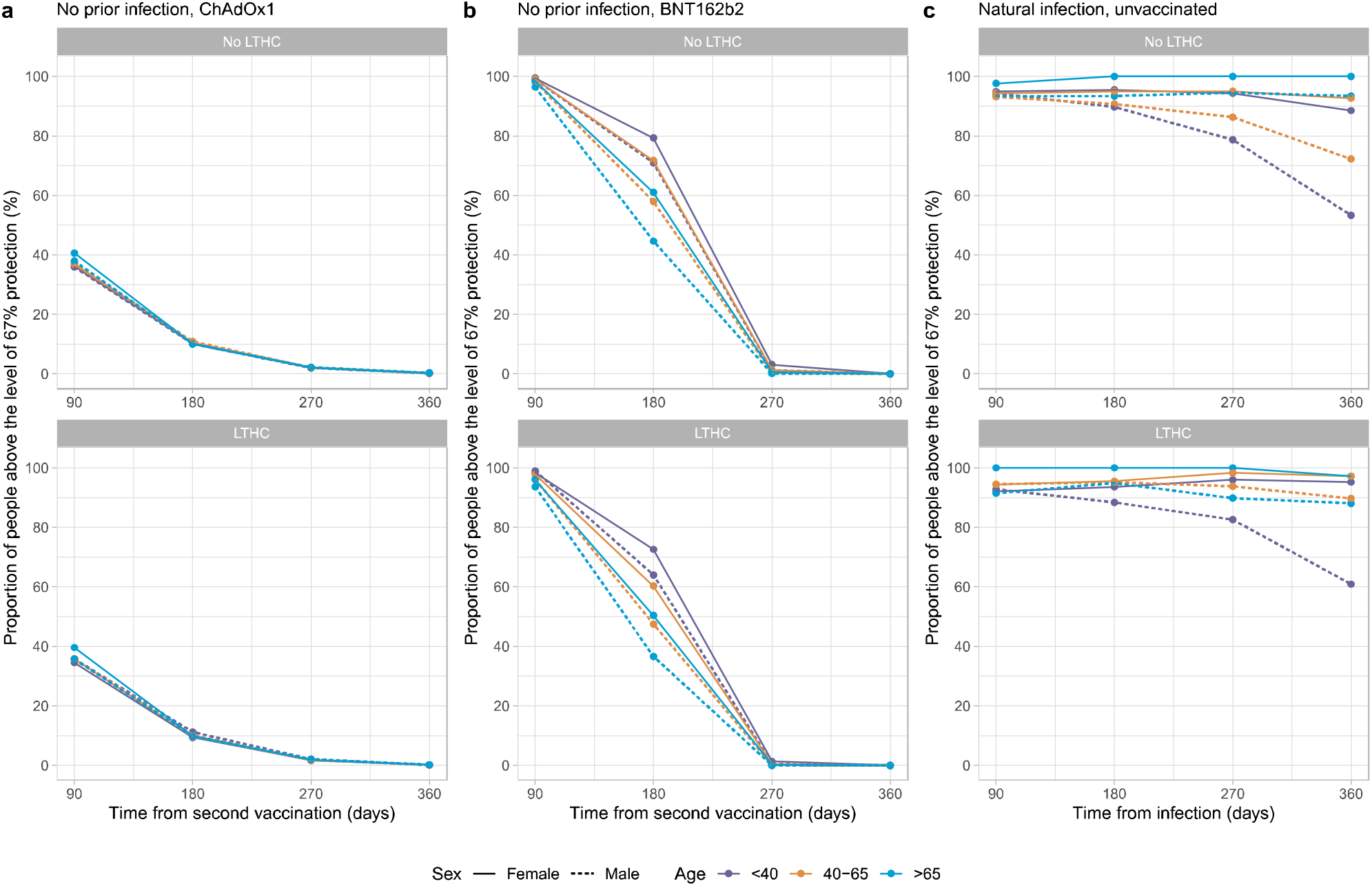
Proportion of participants above the threshold level associated with 67% protection (ChAdOx1: 107 BAU/mL, BNT162b2: 94 BAU/mL, unvaccinated: 33 BAU/mL) by time from second vaccination/infection. **a**, In participants without prior infection and vaccinated with ChAdOX1. **b**, In participants without prior infection and vaccinated with BNT162b2. **c**, In participants who had natural infection and unvaccinated. Estimates were separated by age, sex, and long-term health condition (LTHC). Numbers of participants in each panel were (in the order of no LTHC, LTHC. Numbers in bracket represents <40, 40-65, >65): **a**, N=72,121 [4,973, 46,160, 20,988], 28,518 [2,166, 14,315, 12,037]; **b**, N=36,662 [6,650, 14,108, 15,904], 18,391 [1,800, 6,696, 9,895]; **c**, N=2,625 [1,207, 1,243, 175], 646 [194, 322, 130]. All panels are conditional on participants having antibody response/seroconverting (see section of “Vaccine non-responders” for ChAdOx1 and BNT162b2, and discussion in a previous publication^16^ for unvaccinated individuals).

Emerging viral variants may need higher antibody levels for the same level of neutralising activity^19^; in a sensitivity analysis assuming 2-to 8-fold increases above the threshold associated with 67% protection against the Delta variant, protection from two vaccine doses was short-lived. For example, in a 40-year-old female without any long-term health conditions and without prior infection, if 3-fold higher antibody levels were required, BNT162b2 would provide around 120 days of protection, while ChAdOx1 did not reach the required antibody level; further, if 8-fold higher antibody levels were required, the duration of protection from BNT162b2 would be reduced to only 40 days (**Extended Data Fig. 8**).

### Vaccine non-responders

We previously used latent class mixed models to identify 5.8% and 5.1% of a smaller population of participants receiving one ChAdOx1 or BNT162b2 dose, respectively, as non-responders^20^. Because latent class models would not fit with larger numbers, we used a heuristic rule based on these previous observations to define non-response as all antibody measurements <16 BAU/ml (similar levels to the previous non-response class) and having at least one antibody measurement 21 days after the first or second dose. To examine robustness, we also restricted to those having at least two antibody measurements, and after both doses (rather than each separately). Across different assumptions **(Supplementary Table 6)**, we found that 5.8-7.7% and 3.5-6.0% of participants were classed as non-responders to the first ChAdOx1 or BNT162b2 dose, respectively, similar to previously. Non-responders were older, had a higher percentage of males and white ethnicity, were more deprived, less likely to be working in healthcare, and more likely to have long-term health conditions **(Supplementary Table 7)**. However, only 0.4-1.0% and 0.1-0.5% participants were non-responders to the second ChAdOx1 or BNT162b2 dose, respectively, and 0.3-0.5% and 0.1-0.2% were non-responders to both first and second doses of ChAdOx1 or BNT162b2, respectively.

## Discussion

Based on a large random sample of the UK population, we found significant boosting of anti-spike IgG following second doses of both ChAdOx1 and BNT162b2 vaccines in all age groups and using different dosing intervals, including the three-week dosing interval for BNT162b2. Consistent with our previous findings^20^, those receiving BNT162b2 had significantly higher peak anti-spike IgG responses than ChAdOx1, however, their antibody levels fell faster than following ChAdOx1. After the second vaccination, older age, male sex, and long-term health conditions were all associated with substantially lower peak levels in participants who received BNT162b2 but had smaller effects on peak levels with ChAdOx1. Participants with prior infection had significantly higher peak levels for both vaccines, and their antibody levels fell more slowly. Based on our estimates of anti-spike antibody levels as correlates of protection, antibody levels associated with 67% protection against infection with Delta last for 2-3 months following a second dose of ChAdOx1 and 5-8 months after BNT162b2 in those without prior infection, and could last for 1-2 years in those unvaccinated, but seroconverting following natural infection. Similar to our previous findings^20^, around 6-8% and 4-6% of participants were non/low-responders who did not substantially increase their antibody levels after the first ChAdOx1 or BNT162b2 dose. However, non-response to a second dose of ChAdOx1 or BNT162b2 was much smaller, <1%, suggesting that second doses can significantly boost an initial suboptimal response in most individuals.

Antibody trajectories after vaccination differed substantially by prior infection status. From the Bayesian linear mixed model, prior infection was associated with a significantly higher peak level and a longer half-life for both ChAdOX1 and BNT162b2. Our results are consistent with recent studies in healthcare workers showing that prior SARS-CoV-2 infection leads to higher antibody levels and slower waning post second BNT162b2 vaccination^21–24^. Previous studies also reported that a single dose of BNT162b2 or Sputnik V, an adenovirus-based vaccine, elicited post-vaccination antibody levels that were similar to, or higher than, those without prior infection who received two doses^25–28^. From the generalised additive models, we found slightly lower IgG levels after a single BNT162b2 dose in previously infected participants vs. those with two BNT162b2 doses without prior infection, particularly at older ages. This suggests that a second BNT162b2 dose may still be helpful for previously infected individuals where supplies are sufficient, especially for older age groups. However, for ChAdOx1, the post-second dose IgG levels in those without prior infection were lower than in those previously infected with one ChAdOx1 dose.

In an adjusted Bayesian linear mixed model, longer dosing intervals between 8 and 12 weeks resulted in higher peak antibody levels for ChAdOx1 (6 BAU/mL per week) and BNT162b2 (12 BAU/mL per week), but had no effect on the half-life. Consistent with this, the 3-week dosing interval for BNT162b resulted in a lower peak level compared to an 8-week interval. Other studies also reported lower antibody levels at 2-3 weeks^7^, 4 weeks^8^ and 14-34 days^9^ post-second BNT162b2 dose with 3-week vs extended dosing intervals. However, these studies only measured antibody levels at specific time points after second vaccination, which may not be optimal for comparison given we found that antibody levels were still increasing from 3-6 weeks post-second dose with the 3-week interval (**Extended Data Fig. 2**). We found a slightly longer half-life with 3-week vs. extended dosing intervals, which led to similar duration of protection against new infections despite the lower peak levels.

Older individuals had lower post-second dose IgG peak levels following BNT162b2, but age-related differences were smaller for ChAdOx1. Females had a higher peak IgG level for both vaccines, consistent with widely reported enhanced immune responses in females^29–33^. Healthcare workers had higher IgG peak levels for both vaccines, potentially reflecting a “healthy worker” effect^34^, ongoing occupational exposure, or undetected prior infection. We also found that those reporting non-white ethnicity had higher IgG peak levels. Non-white ethnicity has also been previously associated with higher antibody levels after natural infection^16,35,36^; these findings could be due to genetic or societal differences, or differential rates of undetected prior infection. Long-term health conditions were associated with lower peak levels and shorter half-lives for both vaccines. Our results on BNT162b2 were consistent with a recent large-scale study from Israel, where older individuals, males, and those reporting health conditions had significantly lower IgG levels^37^. Differences were also seen between the two vaccines, e.g. ChAdOx1 had a lower peak level but BNT162b2 had a shorter half-life, and factors generally had larger effects on BNT162b2 than ChAdOx1. Differences in vaccine response are expected given the differing design and mechanism of action of BNT162b2, an mRNA vaccine, and ChAdOx1, an adenovirus vector-based vaccine^38,39^. Alternative booster vaccines may reduce vaccine-specific differences^40^.

We estimated anti-spike IgG mean half-lives after second ChAdOx1 or BNT162b2 doses of 81 days and 52 days, respectively, in those without prior infection, and 94 days and 63 days in those vaccinated and previously infected. The half-life of BNT162b2 was consistent with estimates from previous studies at around 50 days^41–43^. Data from ChAdOx1 and mRNA-1273 vaccine trials showed higher levels of binding and neutralizing antibodies at a limited number of timepoints were associated with a lower risk of infection and a higher vaccine efficacy^14,15^. However, correlating time-updated antibody measurements to protection from infection is important to inform the timing of boosters and other control measures, but existing studies combining correlates of protection and longitudinal data from the same assay are limited. We found higher anti-spike IgG levels were associated with increased protection from infection, but that the level of protection associated with a given antibody level depended on the mechanism generating the antibodies, with natural infection resulting in lower measured antibody levels, but greater protection at a given antibody level, compared to vaccination. We did not see evidence of vaccine-specific differences, with the greater protection from BNT162b vs. ChAdOx1 explained by higher antibody levels rather than increased protection at a given antibody level. Using 67% protection against infection as an example threshold, protection was relatively short-lived in those not previously infected and receiving ChAdOx1; mean levels fell below this threshold at 50-90 days post second dose, but protection was more prolonged following two doses of BNT162b, 160-230 days. In those without prior infection, 6 months after the second dose, only 10% of those with ChAdOx1 would maintain 67% protection, while 40-80% those with BNT162b2 would still be above the threshold, indicating that a booster dose may be prioritised to those who had two doses of ChAdOx1, and would need to be individualised to optimise protection after two BNT162b2 doses. However, 9 months after the second dose of either vaccine, almost everyone without prior infection was below the threshold level associated with 67% protection. Our results are consistent with several recent studies reporting waning vaccine effectiveness 3-6 months after second BNT162b2 or ChAdOx1 vaccinations^44–48^. Estimated durations of protection were much longer following natural infection, ∼1-2 years for those unvaccinated. The protection in those vaccinated with prior infection could be even longer given they have a longer half-life; however, we were not able to quantify this due to limited data. Our estimates of correlates of protection were based on Delta variant infections, and so account for the increased antibody levels needed for neutralisation of this compared with earlier variants^49,50^. However, further increases in antibody levels required for protection with other variants, such as Omicron, would substantially reduce the proportion of the population protected from infection. For example, one study found that neutralisation of Omicron was 8-fold lower than Delta following two BNT162b2 vaccinations^51^; in the case relationship between antibody levels and levels of protection have not changed with Omicron, this would mean antibody levels after two vaccinations would not provide effective protection against Omicron infection **(Extended Data Fig. 8)**. Nevertheless, protection against severe infection is likely to last considerably longer and be potentially more robust^52^. Further long-term follow up data are essential for ongoing monitoring, as it is difficult to predict the emergence of and level of immune protection required for new variants.

Limitations include the fact that we assumed that associations between protection and antibody levels were constant over time, in order to predict vaccine effectiveness from antibody levels beyond the study follow-up. Even though antibody levels decrease over time, other vaccine-induced immune mechanisms, including memory responses, may theoretically provide greater protection at lower antibody levels. There may also be differences in antibody levels and classes in peripheral blood compared to potential exposure sites such as the respiratory tract. Hence, our estimated duration of protection could potentially be underestimated if other immune responses confer longer protection. However, our observations based on waning antibodies are consistent with waning of vaccine effectiveness in our cohort^12^ and other studies^44–48^.

Given the scale of the study, we did not measure other immune responses, including memory-based responses, T cell or innate immune responses which are also involved in protection against infection;^38^ they may explain the greater protection afforded by natural infection than vaccination at the same antibody level. Other study limitations include insufficient data to model two Moderna mRNA-1273 vaccine doses. We measured anti-spike IgG antibody using a single assay, with the upper limit of quantification reached by 13% and 52% of measurements following second ChAdOx1 or BNT162b2 vaccination, respectively, potentially leading to under-estimating peak levels and over-estimating half-lives in those with the highest responses, e.g. younger age groups. However, we used interval censored regression models to address this and capture any waning above the threshold, making it possible to estimate a higher peak level than the upper limit and a more accurate half-life. Neutralising antibodies were not measured, but neutralisation titres were strongly correlated with anti-spike IgG titres **(Extended Data Fig. 9a)**. We also calibrated antibody levels to WHO BAU/mL units for comparison with other studies (**Extended Data Fig. 9b**).

In summary, the second ChAdOx1 or BNT162b2 dose significantly boosts anti-spike IgG levels, and dosing interval has a limited impact on antibody response. Peak levels were higher after BNT162b than ChAdOx1, but older individuals, males, and those with long-term health conditions have lower antibody levels with BNT162b2. Protection based on the threshold level associated with 67% protection can last for 2-3 months for ChAdOx1 and 5-8 months for BNT162b2 in those without prior infection, and can be 1-2 years for those unvaccinated who seroconvert after natural infection. Those vaccinated with prior infection could be protected for >1 year. These results may inform vaccination strategies; a third boosting dose should be prioritised to ChAdOx1 recipients, groups with faster antibody declines, and more clinically vulnerable individuals.

## Methods

### Population and setting

The UK’s Office for National Statistics (ONS) COVID-19 Infection Survey (CIS) (ISRCTN21086382) randomly selects private households on a continuous basis from address lists and previous surveys to provide a representative sample across its four countries (England, Wales, Northern Ireland, Scotland). After obtaining verbal agreement to participate, a study worker visited each household to take written informed consent from individuals ≥2 years. At the first visit, participants were asked for consent for optional follow-up visits every week for the next month, then monthly for 12 months or to April 2022. This consent was obtained from parents/carers for those 2-15 years, while those 10-15 years also provided written assent. Children aged <2 years were not eligible for the study. For the current analysis, we only included participants aged ≥16 years who were eligible for vaccination for the majority of the study period.

Individuals were surveyed on their socio-demographic characteristics, behaviours, and vaccination status. Combined nose and throat swabs were taken from all consenting household members for SARS-CoV-2 PCR testing. For a random 10-20% of households, individuals ≥16 years were invited to provide blood samples monthly for serological testing. Household members of participants who tested positive were also invited to provide blood monthly for follow-up visits. Details on the sampling design are provided elsewhere^53^. From April 2021, additional participants were invited to provide blood samples monthly to assess vaccine responses, based on a combination of random selection and prioritisation of those in the study for the longest period (independent of test results). The study protocol is available at https://www.ndm.ox.ac.uk/covid-19/covid-19-infection-survey/protocol-and-information-sheets. The study received ethical approval from the South Central Berkshire B Research Ethics Committee (20/SC/0195).

From 8^th^ December 2020 to 4^th^ October 2021, 402,348 participants ≥16 years received two ChAdOx1 or BNT162b2 vaccinations. The median (IQR) age was 56 (41-68) years, 216,301 (53.8%) were females, and 375,880 (93.4%) reported white ethnicity. The median (IQR) deprivation percentile was 62 (38-82), and dosing interval was 74 (63-78) days. 11,565 (2.9%) reported working in patient-facing healthcare, and 112,074 (27.9%) reported having a long-term health condition. Among those 402,348 participants, following the design described above with restricted blood sampling, 222,493 participants gave at least one blood sample for antibody testing from 91 days before the first vaccination onwards and were included in our analyses. Participant characteristics were similar between those who did not give blood samples and those who gave blood samples for antibody measurements, and were similar across participants with different number of antibody measurements (1-2, 3-4, and ≥5) (**Supplementary Table 8**).

### Vaccination data

Vaccination information was obtained from participants at visits by self-report, including vaccination type, number of doses, and vaccination dates. Participants from England were also linked to the National Immunisation Management Service (NIMS), which contains all individuals’ vaccination data in the English National Health Service COVID-19 vaccination programme. There was good agreement between self-reported and administrative vaccination data (98% on type and 95% on date^18^). We used vaccination data from NIMS where available for participants from England, and otherwise data from the survey.

Participants aged ≥16 years who received two doses of ChAdOx1 or BNT162b2 from 8^th^ December 2020 onwards with antibody measurements from 91 days before the first vaccination date up until 4^th^ October 2021 were included in the main analysis. Only 4,219 participants received two doses of mRNA-1273 thus were not included **(Extended Data Fig. 1)**.

### Laboratory testing

SARS-CoV-2 antibody levels were measured on venous or capillary blood samples using an ELISA detecting anti-trimeric spike IgG developed by the University of Oxford^53,54^. Normalised results are reported in ng/ml of mAb45 monoclonal antibody equivalents. Before 26 February 2021, the assay used fluorescence detection as previously described, with a positivity threshold of 8 million units validated on banks of known SARS-CoV-2 positive and negative samples^54^. After this, it used a commercialised CE-marked version of the assay, the Thermo Fisher OmniPATH 384 Combi SARS-CoV-2 IgG ELISA (Thermo Fisher Scientific), with the same antigen and colorimetric detection. mAb45 is the manufacturer-provided monoclonal antibody calibrant for this quantitative assay. To allow conversion of fluorometrically determined values in arbitrary units, we compared 3,840 samples which were run in parallel on both systems. A piece-wise linear regression was used to generate the following conversion formula:

log10(mAb45 units) = 0.221738 + 1.751889e-07*fluorescence_units + 5.416675e-07*(fluorescence_units>9190310)*(fluorescence_units-9190310)

We calibrated the results of the Thermo Fisher OmniPATH assay into WHO international units (binding antibody unit, BAU/mL) using serial dilutions of National Institute for Biological Standards and Control (NIBSC) Working Standard 21/234. The NIBSC 21/234 Working Standard has been previously calibrated against the WHO International Standard for anti-SARS-CoV-2 immunoglobulin (NIBSC code 20/136), with anti-spike IgG potency of 832 BAU/mL (95%CI 746-929). We generated 2-fold dilutions of 21/234 between 1:400 and 1:8000 from three separate batches on three separate days. Results from a total of 63 diluted samples were merged and a linear regression model fitted constrained to have an intercept of zero to convert mAB45 units in ng/ml for samples diluted at 1:50 to BAU/mL (**Extended Data Fig. 9b**):

BAU/mL = 0.559 * [mAb45 concentration in ng/mL at 1:50]

We used ≥23 BAU/mL as the threshold for determining IgG positivity (corresponding to the 8 million units with fluorescence detection). Given the lower and upper limits of the assay, measurements <1 BAU/mL (2,922 observations, 0.4%) and >450 BAU/mL (146,337 observations, 19.2%) were truncated at 1 and 450 BAU/mL, respectively.

Combined nose and throat swabs were tested by PCR assays using the Thermo Fisher TaqPath SARS-CoV-2 assay at high-throughput national ‘Lighthouse’ laboratories in Glasgow and Milton Keynes (up until 8 February 2021). PCR outputs were analysed using UgenTec FastFinder 3.300.5, with an assay-specific algorithm and decision mechanism that allows conversion of amplification assay raw data into test results with minimal manual intervention. Positive samples are defined as having at least a single N-gene and/or ORF1ab detected (although S-gene cycle threshold (Ct) values are determined, S-gene detection alone is not considered sufficient to call a sample positive^53^) and PCR traces exhibiting an appropriate morphology.

### Statistical analysis

Analysis of antibody levels included participants aged ≥16 years who received two doses of ChAdOx1 or BNT162b2 vaccines with or without prior SARS-CoV-2 infection. Age was truncated at 85 years in all analyses to reduce the influence of outliers. Prior infection was defined as having a PCR-positive swab test recorded in the survey or the English national testing programme (national testing data were not available for Scotland, Wales, and Northern Ireland), or a prior positive anti-spike IgG result (≥23 BAU/mL) any time before the first vaccination. Where participants were known to have become infected after vaccination (based on positive PCR tests from the survey or national linked data), antibody measured after infection were excluded from the analyses. The dosing interval was calculated from the first and second vaccination dates. For the main analysis, we excluded a small number of participants who were considered as non-responders after the first or second dose, which was defined as all antibody measurements being <16 BAU/mL and having at least one antibody measurement 21 days after the first or second dose (N=5,098 excluded for ChAdOx1, N=1,649 excluded for BNT162b2) **(Extended Data Fig. 1)**. We also excluded participants with recorded dosing interval <49 days or >91 days for ChAdOX1 (N=4,748 excluded), and 29-48 days or >91 days for BNT162b2 (N=6,374 excluded). 17-28 days were classified as a 3-week interval for BNT162b2.

We used linear generalized additive models (GAMs) to model anti-spike IgG antibody measurements after the first and second dose in order to identify the most appropriate timepoints to model antibody declines post second vaccination in more detail. Antibody measurements truncated at 450 BAU/mL were counted as 450 BAU/mL. We built separate models by vaccine type and prior infection status given the hypothesis that antibody response would vary by these two factors. Each model was minimally adjusted for only age and dosing interval using a tensor product of B-splines to allow for non-linearity and interaction among age, dosing interval, and time since vaccination, setting the date of the second vaccination as t=0. The smoothing penalty was selected using fast restricted maximum likelihood as implemented in the mcgv R package. The 95% CIs were calculated using the following formula: prediction ± 1.96 × standard error of prediction. We only included antibody measurements from 14 days before the first dose (setting the most recent measurement prior to 14 days before the first dose as 14 days) for those with no evidence of prior infection, and excluded measurements taken after the 95^th^ percentile of the observed t>0 time points to avoid the outlier influence.

We used Bayesian linear mixed interval-censored models to estimate changes in antibody levels after the second ChAdOx1 or BNT162b2 dose. We included measurements from 21 days post-second dose reflecting the peak level from the GAMs (except for 3-week BNT162b2, see below). Measurements taken after the 95^th^ percentile of the observed time points from 21 days post-second dose were excluded to avoid outlier influence. We assumed an exponential fall in antibody levels over time, i.e., a linear decline on a log2 scale. To examine non-linearity in antibody declines, especially the assumption that the rate of antibody decline would flatten, we additionally fitted a model using 4-knot splines for time (knots placed at 10^th^, 40^th^, 60^th^, and 90^th^ of observed time points) and compared the model fit with the log-linear model using the leave-one-out cross-validation information criterion (LOOIC). We found that the spline model had a higher LOOIC (indicating a worse model fit) than the log-linear model for ChAdOx1 (382954 vs 378640) and BNT162b2 (252730 vs 240472). For both vaccines, the estimated trajectories were similar and there was no evidence of antibody decline flattening (**Extended Data Fig. 10**), so we used the log-linear model for the rest of the analysis.

Population-level fixed effects, individual-level random effects for intercept and slope, and correlation between random effects were included in both models. The outcome was right-censored at 450 BAU/mL reflecting truncation of IgG values at the upper limit of quantification (i.e. all measurements truncated to 450 BAU/mL were considered to be >450 BAU/mL in analyses). We built a multivariable model to examine the association between peak levels and antibody half-lives with continuous age (16-85 years), sex, ethnicity, report having a long-term health condition, report working in patient-facing healthcare, deprivation percentile, continuous dosing interval (7-13 weeks), and prior infection status for both vaccines. For BNT162b2, we additionally examined the impact of a 3-week dosing interval (17-28 days) by creating a binary variable and excluding antibody measurements ≤42 days post-second dose for the 3-week group (identified from the GAM as they peaked at around 42 days post-second dose).

For each Bayesian linear mixed interval-censored model, weakly informative priors were used **(Supplementary Table 9)**. Four chains were run per model with 4,000 iterations and a warm-up period of 2,000 iterations to ensure convergence, which was confirmed visually and by ensuring the Gelman-Rubin statistic was <1.05 **(Supplementary Table 10)**. 95% credible intervals were calculated using highest posterior density intervals.

For the analysis of correlates of protection, we used data from study visits from 17^th^ May 2021 to 4^th^ October 2021. These visits were from 35-298 days after the first vaccination, which captured the time period over which real-world vaccine effectiveness has been observed to wane (**Supplementary Fig. 4** and **Supplementary Table 5**). We grouped positive tests into episodes because PCR-positive results might be observed at multiple visits after infection. Following previous work, we defined the start of a new episode or ‘positive case’ as the date of (1) the first PCR-positive test in the study (not preceded by any study PCR-positive test); (2) a PCR-positive test after four or more consecutive negative visits; or (3) a PCR-positive test at least 120 days after the start of a previous episode with one or more negative tests immediately preceding this^12^. Analyses were based on visits, dropping any visits where participants were not at risk due to a recent new positive PCR test, with new PCR-positive episodes as the outcome. We used separate logistic generalised additive models for three outcomes: any positive PCR episode, a positive PCR episode with a moderate to high viral load (Ct value <30), a positive PCR episode with self-reported symptoms. Two exposure specifications were investigated. Firstly, considering the effect of the most recent antibody measurement obtained 21-59 days prior to the current visit, excluding more recent measurements to avoid changes in antibody levels arising from recent infection that might only be detected at the routine study visit despite occurring before this. Alternatively, we considered the maximum antibody measurement obtained ≥21 days prior to the visit. The relationship between antibody levels and the outcome was modelled using thin plate splines. We adjusted for the following confounders in all models: geographic area (12 regions in England, or Wales, Scotland or Northern Ireland) and age in years, rural-urban home address, sex, ethnicity (white vs. non-white), household size, multigenerational household, deprivation, presence of long-term health conditions, working in a care-home, having a patient-facing role in health or social care, direct or indirect contact with a hospital or care-home, smoking status, and visit frequency. Calendar time and age were included using a tensor spline which was allowed to vary by region/country^12^.

Three groups were investigated, unvaccinated participants with or without evidence of prior infection (to assess the impact of prior infection), participants vaccinated with ChAdOx1, and participants vaccinated with BNT162b2. Vaccinated participants with evidence of prior infection were excluded, as there were insufficient data to model these groups separately and the relationship between antibody levels and protection may differ in these groups. Visits occurring in the 21 days prior to vaccination were excluded, as we have previously reported infection rates change in the run-up to vaccination^18^. Visits in vaccinated participants were included from ≥21 days after first vaccination. In secondary analyses we grouped vaccinated individuals into one category.

All analyses were performed in R 3.6 using the following packages: tidyverse (version 1.3.0), mgcv (version 1.8-31), brms (version 2.14.0), rstanarm (version 2.21.1), splines (version 3.6.1), ggeffects (version 0.14.3), arsenal (version 3.4.0), cowplot (version 1.1.0), bayesplot (version 1.7.2).

## Supporting information

supplementary results

## Data Availability

Data are still being collected for the COVID-19 Infection Survey. De-identified study data are available for access by accredited researchers in the ONS Secure Research Service (SRS) for accredited research purposes under part 5, chapter 5 of the Digital Economy Act 2017. For further information about accreditation, contact or visit the SRS website.

## Acknowledgements

We are grateful for the support of all COVID-19 Infection Survey participants.

This study is funded by the Department of Health and Social Care with in-kind support from the Welsh Government, the Department of Health on behalf of the Northern Ireland Government and the Scottish Government. JW is supported by University of Oxford and the China Scholarship Council. ASW, TEAP, NS, DE, KBP are supported by the National Institute for Health Research Health Protection Research Unit (NIHR HPRU) in Healthcare Associated Infections and Antimicrobial Resistance at the University of Oxford in partnership with the UK Health Security Agency (UK HSA) (NIHR200915). ASW and TEAP are also supported by the NIHR Oxford Biomedical Research Centre. KBP is also supported by the Huo Family Foundation. ASW is also supported by core support from the Medical Research Council UK to the MRC Clinical Trials Unit [MC_UU_12023/22] and is an NIHR Senior Investigator. PCM is funded by Wellcome (intermediate fellowship, grant ref 110110/Z/15/Z) and holds an NIHR Oxford BRC Senior Fellowship award. DWE is supported by a Robertson Fellowship and an NIHR Oxford BRC Senior Fellowship. NS is an Oxford Martin Fellow and holds an NIHR Oxford BRC Senior Fellowship. The views expressed are those of the authors and not necessarily those of the National Health Service, NIHR, Department of Health, or UKHSA.

## COVID-19 Infection Survey team group authorship

**Office for National Statistics**: Sir Ian Diamond, Emma Rourke, Ruth Studley, Tina Thomas, Duncan Cook.

**Office for National Statistics COVID Infection Survey Analysis and Operations teams**, in particular Daniel Ayoubkhani, Russell Black, Antonio Felton, Megan Crees, Joel Jones, Lina Lloyd, Esther Sutherland.

**University of Oxford, Nuffield Department of Medicine**: Ann Sarah Walker, Derrick Crook, Philippa C Matthews, Tim Peto, Emma Pritchard, Nicole Stoesser, Karina-Doris Vihta, Jia Wei, Alison Howarth, George Doherty, James Kavanagh, Kevin K Chau, Stephanie B Hatch, Daniel Ebner, Lucas Martins Ferreira, Thomas Christott, Brian D Marsden, Wanwisa Dejnirattisai, Juthathip Mongkolsapaya, Sarah Cameron, Phoebe Tamblin-Hopper, Magda Wolna, Rachael Brown, Sarah Hoosdally, Richard Cornall, David I Stuart, Gavin Screaton.

**University of Oxford, Nuffield Department of Population Health**: Koen Pouwels.

**University of Oxford, Big Data Institute:** David W Eyre, Katrina Lythgoe, David Bonsall, Tanya Golubchik, Helen Fryer.

**University of Oxford, Radcliffe Department of Medicine**: John Bell.

**Oxford University Hospitals NHS Foundation Trust:** Stuart Cox, Kevin Paddon, Tim James.

**University of Manchester**: Thomas House.

**Public Health England**: John Newton, Julie Robotham, Paul Birrell.

**IQVIA**: Helena Jordan, Tim Sheppard, Graham Athey, Dan Moody, Leigh Curry, Pamela Brereton.

**National Biocentre**: Ian Jarvis, Anna Godsmark, George Morris, Bobby Mallick, Phil Eeles.

**Glasgow Lighthouse Laboratory**: Jodie Hay, Harper VanSteenhouse.

**Department of Health and Social Care**: Jessica Lee.

**Welsh Government**: Sean White, Tim Evans, Lisa Bloemberg.

**Scottish Government**: Katie Allison, Anouska Pandya, Sophie Davis.

**Public Health Scotland**: David I Conway, Margaret MacLeod, Chris Cunningham.

## Author Contributions

The study was designed and planned by ASW, JF, JB, JN, ID and KBP, and is being conducted by ASW, RS, DC, ER, AH, BM, TEAP, PCM, NS, SH, EYJ, DIS, DWC and DWE. This specific analysis was designed by JW, DWE, ASW, and KBP. JW and KBP contributed to the statistical analysis of the survey data. JW, DWE, KBP and ASW drafted the manuscript and all authors contributed to interpretation of the data and results and revised the manuscript. All authors approved the final version of the manuscript.

## Code availability

A copy of the analysis code is available at https://github.com/jiaweioxford/COVID19_second_vaccine_antibody_response.

## References

1. Medicines and Healthcare products Regulatory Agency. Regulatory approval of Pfizer/BioNTech vaccine for COVID-19 - GOV.UK. https://www.gov.uk/government/publications/regulatory-approval-of-pfizer-biontech-vaccine-for-covid-19 (2020).

2. Medicines and Healthcare products Regulatory Agency. Oxford University/AstraZeneca COVID-19 vaccine approved - GOV.UK. https://www.gov.uk/government/news/oxford-universityastrazeneca-covid-19-vaccine-approved (2020).

3. Department of Health and Social Care. UK COVID-19 vaccines delivery plan Contents. https://www.gov.uk/government/publications/uk-covid-19-vaccines-delivery-plan (2021).

4. Vaccinations in the UK | Coronavirus in the UK. https://coronavirus.data.gov.uk/details/vaccinations (2021).

5. Prioritising the first COVID-19 vaccine dose: JCVI statement - GOV.UK. https://www.gov.uk/government/publications/prioritising-the-first-covid-19-vaccine-dose-jcvi-statement.

6. Voysey, M. et al. Safety and efficacy of the ChAdOx1 nCoV-19 vaccine (AZD1222) against SARS-CoV-2: an interim analysis of four randomised controlled trials in Brazil, South Africa, and the UK. The Lancet 397, 99–111 (2021).

7. Parry, H. et al. Extended interval BNT162b2 vaccination enhances peak antibody generation in older people. medRxiv 2021.05.15.21257017 (2021) doi:10.1101/2021.05.15.21257017.

8. Payne, R. P. et al. Immunogenicity of standard and extended dosing intervals of BNT162b2 mRNA vaccine. Cell 184, 5699-5714.e11 (2021).

9. Amirthalingam, G. et al. Serological responses and vaccine effectiveness for extended COVI-19 vaccine schedules in England. Nature Communications 2021 12:1 12, 1–9 (2021).

10. Sheikh, A., McMenamin, J., Taylor, B. & Robertson, C. SARS-CoV-2 Delta VOC in Scotland: demographics, risk of hospital admission, and vaccine effectiveness. The Lancet 397, 2461–2462 (2021).

11. Bernal, J. L. et al. Effectiveness of Covid-19 Vaccines against the B.1.617.2 (Delta) Variant. New England Journal of Medicine 385, 585–594 (2021).

12. Pouwels, K. B. et al. Effect of Delta variant on viral burden and vaccine effectiveness against new SARS-CoV-2 infections in the UK. Nature Medicine 2021 27:12 27, 2127–2135 (2021).

13. Most vulnerable offered second dose of COVID-19 vaccine earlier to help protect against variants - GOV.UK. https://www.gov.uk/government/news/most-vulnerable-offered-second-dose-of-covid-19-vaccine-earlier-to-help-protect-against-variants (2021).

14. Feng, S. et al. Correlates of protection against symptomatic and asymptomatic SARS-CoV-2 infection. Nature Medicine 2021 27:11 27, 2032–2040 (2021).

15. Gilbert, P. B. et al. Immune correlates analysis of the mRNA-1273 COVID-19 vaccine efficacy clinical trial. Science eab3435 (2021).

16. Wei, J. et al. Anti-spike antibody response to natural SARS-CoV-2 infection in the general population. Nature Communications 2021 12:1 12, 1–12 (2021).

17. SARS-CoV-2 variants of concern and variants under investigation in England. https://assets.publishing.service.gov.uk/government/uploads/system/uploads/attachment_data/file/1025827/Technical_Briefing_25.pdf (2021).

18. Pritchard, E. et al. Impact of vaccination on new SARS-CoV-2 infections in the United Kingdom. Nature Medicine 2021 27:8 27, 1370–1378 (2021).

19. Noori, M. et al. Potency of BNT162b2 and mRNA-1273 vaccine-induced neutralizing antibodies against severe acute respiratory syndrome-CoV-2 variants of concern: A systematic review of in vitro studies. Reviews in Medical Virology e2277 (2021) doi:10.1002/RMV.2277.

20. Wei, J. et al. Antibody responses to SARS-CoV-2 vaccines in 45,965 adults from the general population of the United Kingdom. Nature Microbiology 2021 6:9 6, 1140–1149 (2021).

21. Zhong, D. et al. Durability of Antibody Levels After Vaccination With mRNA SARS-CoV-2 Vaccine in Individuals With or Without Prior Infection. JAMA 326, 2524–2526 (2021).

22. Urbanowicz, R. A. et al. Two doses of the SARS-CoV-2 BNT162b2 vaccine enhance antibody responses to variants in individuals with prior SARS-CoV-2 infection. Science Translational Medicine 13, 847 (2021).

23. Buonfrate, D. et al. Antibody response induced by the BNT162b2 mRNA COVID-19 vaccine in a cohort of health-care workers, with or without prior SARS-CoV-2 infection: a prospective study. Clinical microbiology and infectionr!: the official publication of the European Society of Clinical Microbiology and Infectious Diseases 27, 1845–1850 (2021).

24. Fraley, E. et al. The Impact of Prior Infection and Age on Antibody Persistence After Severe Acute Respiratory Syndrome Coronavirus 2 Messenger RNA Vaccine. Clinical Infectious Diseases (2021) doi:10.1093/CID/CIAB850.

25. Vicenti, I. et al. The second dose of the BNT162b2 mRNA vaccine does not boost SARS-CoV-2 neutralizing antibody response in previously infected subjects. Infection 1, 1 (2021).

26. Krammer, F. et al. Antibody Responses in Seropositive Persons after a Single Dose of SARS-CoV-2 mRNA Vaccine. New England Journal of Medicine 384, 1372–1374 (2021).

27. Ebinger, J. E. et al. Antibody responses to the BNT162b2 mRNA vaccine in individuals previously infected with SARS-CoV-2. Nature Medicine 2021 27:6 27, 981–984 (2021).

28. Claro, F., Silva, D., Rodriguez, M., Rangel, H. R. & de Waard, J. H. Immunoglobulin G antibody response to the Sputnik V vaccine: previous SARS-CoV-2 seropositive individuals may need just one vaccine dose. International Journal of Infectious Diseases 111, 261–266 (2021).

29. Terpos, E. et al. Age-dependent and gender-dependent antibody responses against SARS-CoV-2 in health workers and octogenarians after vaccination with the BNT162b2 mRNA vaccine. American Journal of Hematology 96, E257–E259 (2021).

30. Amodio, E. et al. Antibodies Responses to SARS-CoV-2 in a Large Cohort of Vaccinated Subjects and Seropositive Patients. Vaccines 2021, Vol. 9, Page 714 9, 714 (2021).

31. Ward, H. et al. Vaccine uptake and SARS-CoV-2 antibody prevalence among 207,337 adults during May 2021 in England: REACT-2 study. medRxiv 2021.07.14.21260497 (2021) doi:10.1101/2021.07.14.21260497.

32. Takahashi, T. et al. Sex differences in immune responses that underlie COVID-19 disease outcomes. Nature 588, 315–320 (2020).

33. Bunders, M. J. & Altfeld, M. Implications of Sex Differences in Immunity for SARS-CoV-2 Pathogenesis and Design of Therapeutic Interventions. Immunity vol. 53 487–495 (2020).

34. Li, C.-Y. & Sung, E.-C. A review of the healthy worker effect in occupational epidemiology. Occup. Mod vol. 49 https://academic.oup.com/occmed/article/49/4/225/1387118 (1999).

35. Shields, A. M. et al. Serological responses to SARS-CoV-2 following non-hospitalised infection: clinical and ethnodemographic features associated with the magnitude of the antibody response. BMJ Open Respiratory Research 8, e000872 (2021).

36. Lumley, S. F. et al. The Duration, Dynamics, and Determinants of Severe Acute Respiratory Syndrome Coronavirus 2 (SARS-CoV-2) Antibody Responses in Individual Healthcare Workers. Clinical Infectious Diseases 73, e699–e709 (2021).

37. Levin, E. G. et al. Waning Immune Humoral Response to BNT162b2 Covid-19 Vaccine over 6 Months. New England Journal of Medicine 385, e84 (2021).

38. Sadarangani, M., Marchant, A. & Kollmann, T. R. Immunological mechanisms of vaccine-induced protection against COVID-19 in humans. Nature Reviews Immunology 2021 21:8 21, 475–484 (2021).

39. Tregoning, J. S., Flight, K. E., Higham, S. L., Wang, Z. & Pierce, B. F. Progress of the COVID-19 vaccine effort: viruses, vaccines and variants versus efficacy, effectiveness and escape. Nature Reviews Immunology 2021 21:10 21, 626–636 (2021).

40. Liu, X. et al. Safety and immunogenicity of heterologous versus homologous prime-boost schedules with an adenoviral vectored and mRNA COVID-19 vaccine (Com-COV): a single-blind, randomised, non-inferiority trial. The Lancet 398, 856–869 (2021).

41. Favresse, J. et al. Antibody titres decline 3-month post-vaccination with BNT162b2. Emerging microbes & infections 10, 1495–1498 (2021).

42. van Praet, J. T. et al. Dynamics of the Cellular and Humoral Immune Response After BNT162b2 Messenger Ribonucleic Acid Coronavirus Disease 2019 (COVID-19) Vaccination in COVID-19-Naive Nursing Home Residents. The Journal of Infectious Diseases 224, 1690–1693 (2021).

43. Maeda, K. et al. Correlates of neutralizing/SARS-CoV-2-S1-binding antibody response with adverse effects and immune kinetics in BNT162b2-vaccinated individuals. Scientific Reports 2021 11:1 11, 1–10 (2021).

44. Tartof, S. Y. et al. Effectiveness of mRNA BNT162b2 COVID-19 vaccine up to 6 months in a large integrated health system in the USA: a retrospective cohort study. The Lancet 398, 1407–1416 (2021).

45. Goldberg, Y. et al. Waning Immunity after the BNT162b2 Vaccine in Israel. New England Journal of Medicine 385, e85 (2021).

46. Eyre, D. W. et al. Effect of Covid-19 Vaccination on Transmission of Alpha and Delta Variants. New England Journal of Medicine (2022) doi:10.1056/NEJMOA2116597.

47. Bergwerk, M. et al. Covid-19 Breakthrough Infections in Vaccinated Health Care Workers. New England Journal of Medicine 385, 1474–1484 (2021).

48. SARS-CoV-2 variants of concern and variants under investigation in England. https://assets.publishing.service.gov.uk/government/uploads/system/uploads/attachment_data/file/1044481/Technical-Briefing-31-Dec-2021-Omicron_severity_update.pdf (2021).

49. Jalkanen, P. et al. COVID-19 mRNA vaccine induced antibody responses against three SARS-CoV-2 variants. Nature Communications 2021 12:1 12, 1–11 (2021).

50. Planas, D. et al. Reduced sensitivity of SARS-CoV-2 variant Delta to antibody neutralization. Nature 2021 596:7871 596, 276–280 (2021).

51. Nemet, I. et al. Third BNT162b2 Vaccination Neutralization of SARS-CoV-2 Omicron Infection. The New England journal of medicine (2021) doi:10.1056/NEJMC2119358/SUPPL_FILE/NEJMC2119358_DISCLOSURES.PDF.

52. Khoury, D. S. et al. Neutralizing antibody levels are highly predictive of immune protection from symptomatic SARS-CoV-2 infection. Nature Medicine 2021 27:7 27, 1205–1211 (2021).

53. Pouwels, K. B. et al. Community prevalence of SARS-CoV-2 in England from April to November, 2020: results from the ONS Coronavirus Infection Survey. The Lancet Public Health 6, e30–e38 (2021).

54. Ainsworth, M. et al. Performance characteristics of five immunoassays for SARS-CoV-2: a head-to-head benchmark comparison. The Lancet Infectious Diseases 20, 1390–1400 (2020).

