## supplementary results for "SARS-CoV-2 anti-spike IgG antibody responses after second dose of ChAdOx1 or BNT162b2 and correlates of protection in the UK general population"

### Extended Data Figures

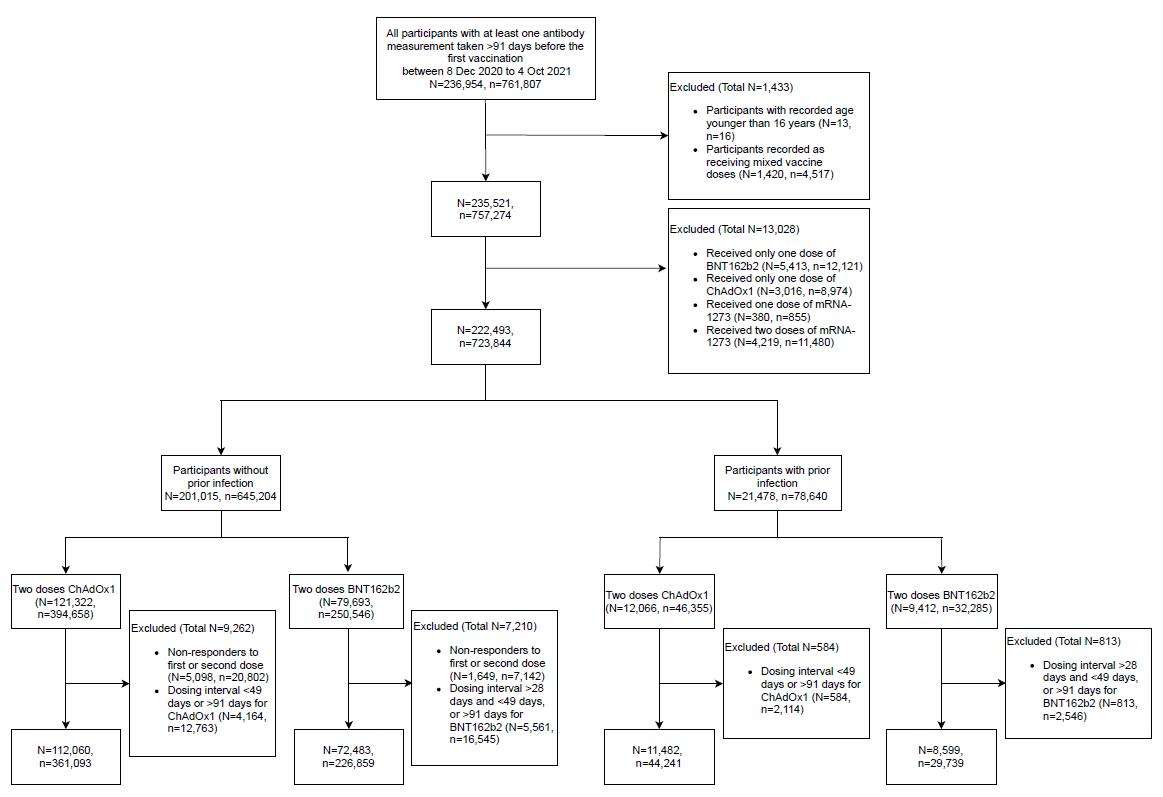

**Extended Data Fig. 1.** **Flowchart of the study cohort.** *N* represents the number of participants, and *n* represents the number of antibody measurements.

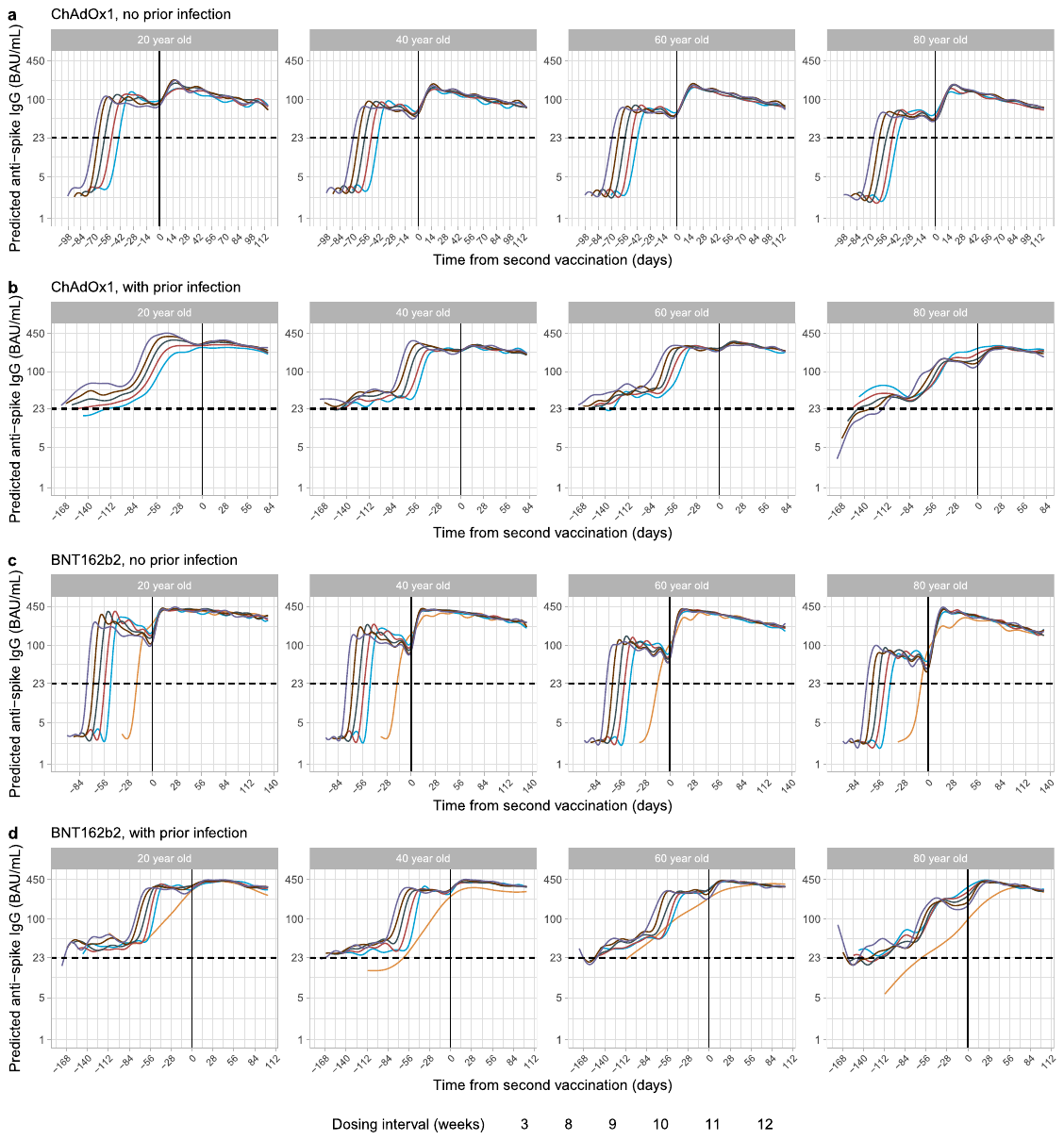

**Extended Data Fig. 2.** **Mean anti-spike IgG levels by time from second vaccination in 20-, 40-, 60- and 80-year-olds according to dosing interval using generalised additive models adjusted for age and dosing interval**. **a,** Participants who received two doses of ChAdOx1 without prior infection. **b,** Participants who received two doses of ChAdOx1 with prior infection. **c**, Participants who received two doses of BNT162b2 without prior infection. **d**, Participants who received two doses of BNT162b2 with prior infection. Different *x* axis scales reflect different durations of follow-up post-vaccination in the different cohorts. Predicted levels are plotted on a log scale. Black dotted line indicates the threshold of IgG positivity (23 BAU/mL). Black solid line indicates the second vaccination date. Line colour indicates response predicted for 3 weeks, 8-12 weeks dosing interval. The 95% CIs are calculated by prediction ± 1.96 × standard error of the prediction.

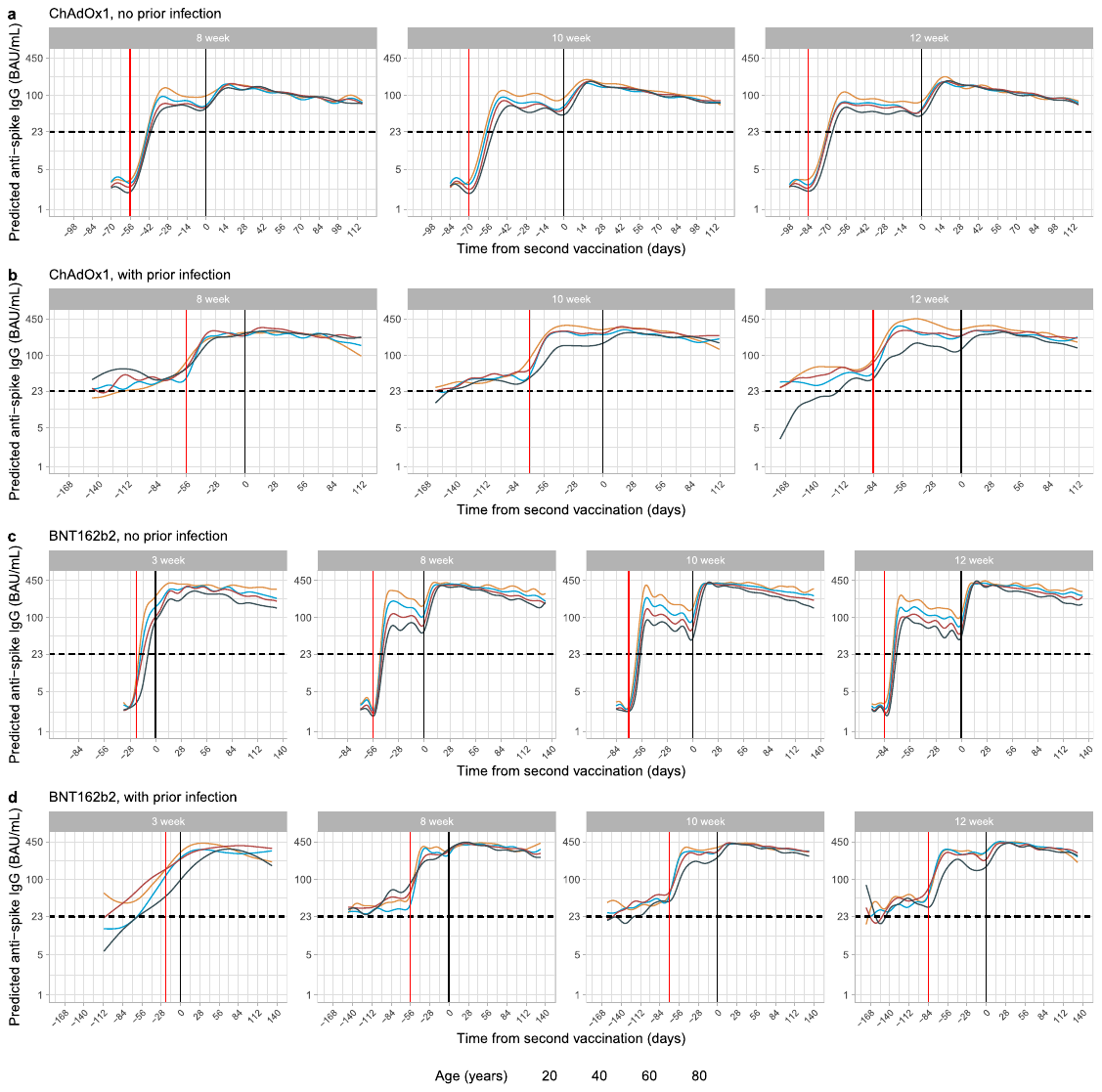

**Extended Data Fig. 3.** **Mean anti-spike IgG levels by time from second vaccination in 3-, 8-, 10-, and 12-week dosing interval groups according to age using generalised additive models adjusted for age and dosing interval.** **a,** Participants who received two doses of ChAdOx1 without prior infection. **b,** Participants who received two doses of ChAdOx1 with prior infection. **c**, Participants who received two doses of BNT162b2 without prior infection. **d**, Participants who received two doses of BNT162b2 with prior infection. Different *x* axis scales reflect different durations of follow-up post-vaccination in the different cohorts. Predicted levels are plotted on a log scale. Black dotted line indicates the threshold of IgG positivity (23 BAU/mL). Red solid line indicates the first vaccination and black solid line indicates the second vaccination. Line colour indicates response predicted for age 20, 40, 60, and 80 years. The 95% CIs are calculated by prediction ± 1.96 × standard error of the prediction.

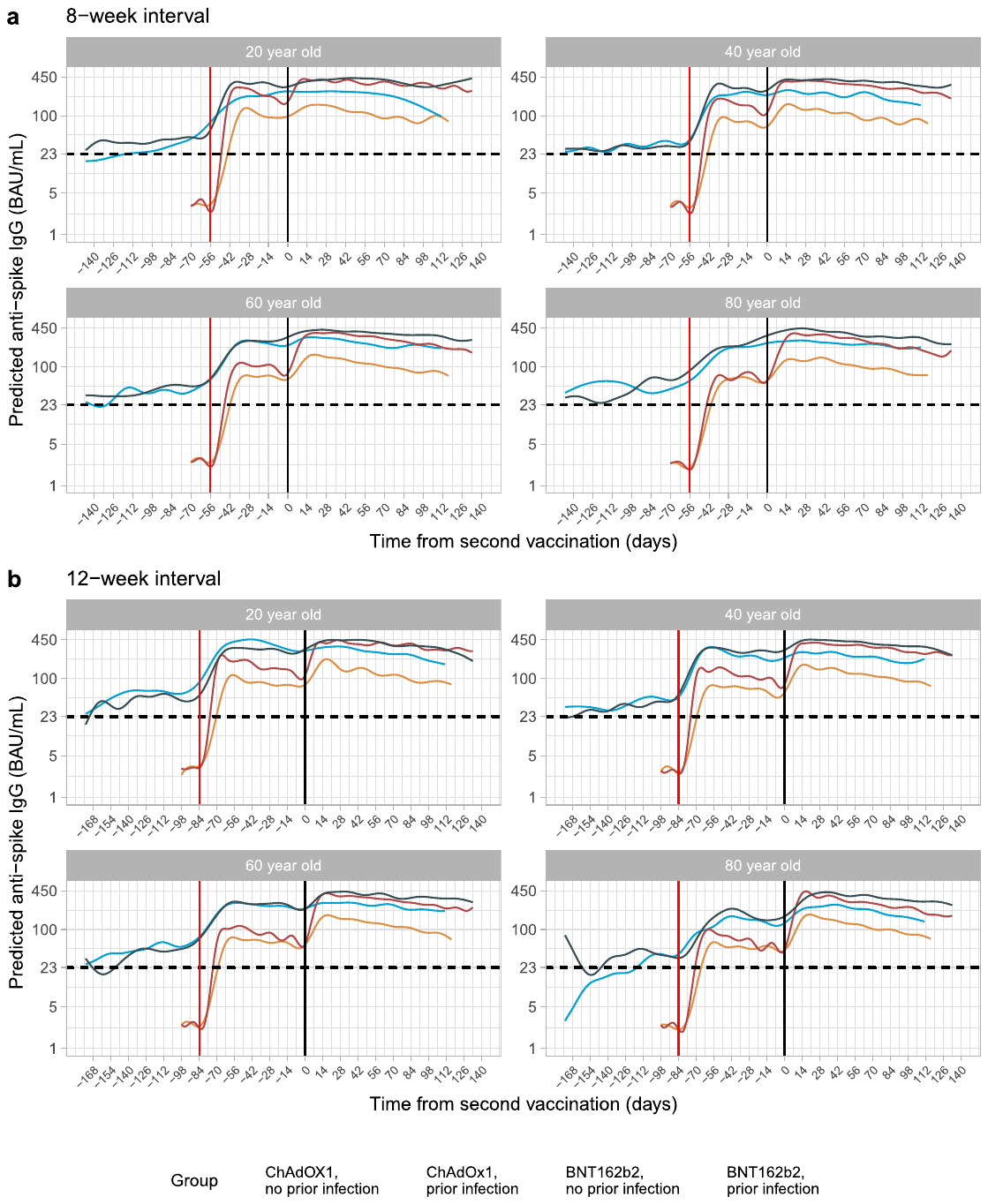

**Extended Data Fig. 4.** **Mean anti-spike IgG levels by time from second vaccination according to vaccine type, prior infection status, and age using generalised additive models adjusted for age and dosing interval.** **a,** Participants with a 8-week dosing interval. **b,** Participants with a 12-week dosing interval. Predicted levels are plotted on a log scale. Black dotted line indicates the threshold of IgG positivity (23 BAU/mL). Red solid line indicates the first vaccination and black solid line indicates the second vaccination. Line colour indicates response predicted for ChAdOx1 and BNT162b2, with or without prior infection. The 95% CIs are calculated by prediction ± 1.96 × standard error of the prediction.

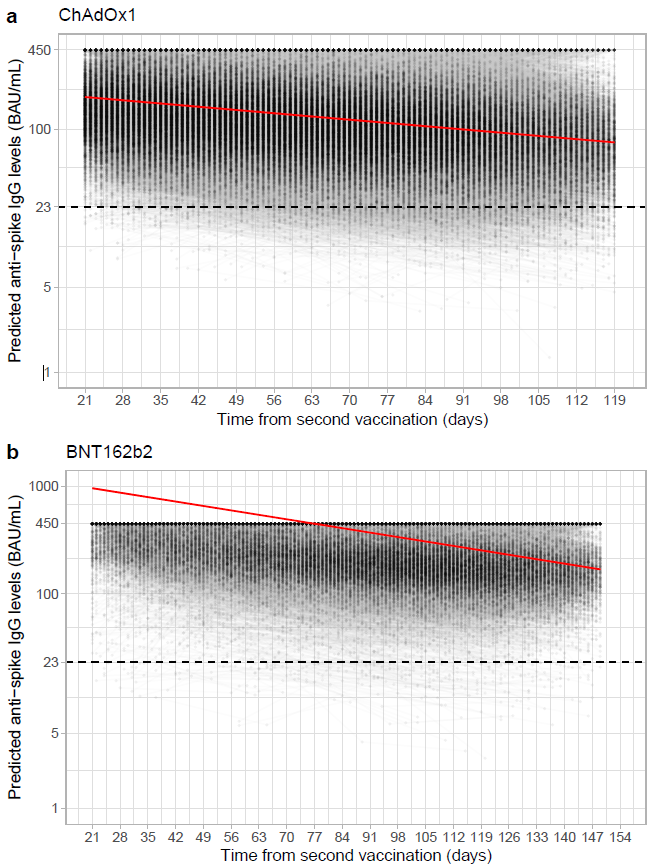

**Extended Data Fig. 5.** **Estimated unadjusted mean trajectory of anti-spike IgG antibody levels and individual trajectories from 21 days after the second vaccination. a,** In participants who received two doses of ChAdOx1. **b,** In participants who received two doses of BNT162b2. Black dashed line indicates the assay threshold for IgG positivity (23 BAU/mL). For BNT162b2, interval censored regression takes account of the fact that 52% measurements were above the upper quantification limit of 450 BAU/mL, and the timing of these measurements, so the estimated peak levels were higher than 450 BAU/mL.

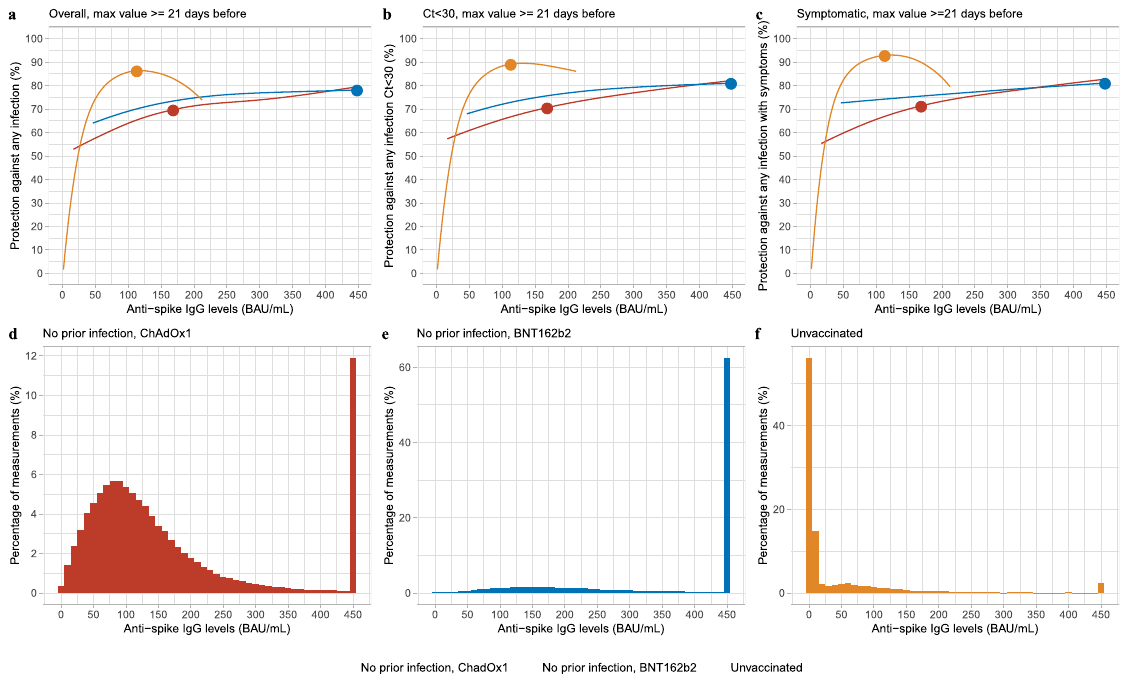

**Extended Data Fig. 6. Association between anti-spike IgG levels and protection from SARS-CoV-2 infection using the maximum antibody measurement obtained ≥21 days prior to the visit. a,** protection against any infection; **b**, protection against infection with a moderate to high viral load (Ct value <30); **c**, protection against infection with self-reported symptoms. Three groups are investigated, unvaccinated participants with or without evidence of prior infection, participants vaccinated with ChAdOx1 without evidence of prior infection, and participants vaccinated with BNT162b2 without evidence of prior infection. Dots represent the median predicted individual peak levels from the Bayesian linear mixed models: 1026 BAU/mL for BNT162b2 (plotted at the upper quantification limit of 450 BAU/mL), 167 BAU/mL for ChAdOx1, and 111 BAU/mL for unvaccinated participants^1^. Distribution of the maximum anti-spike IgG measurements for the three population groups are shown in panel **d, e, f**. See **Supplementary Fig. 4** and **Supplementary Table 5** for timing of visits relative to second vaccination.

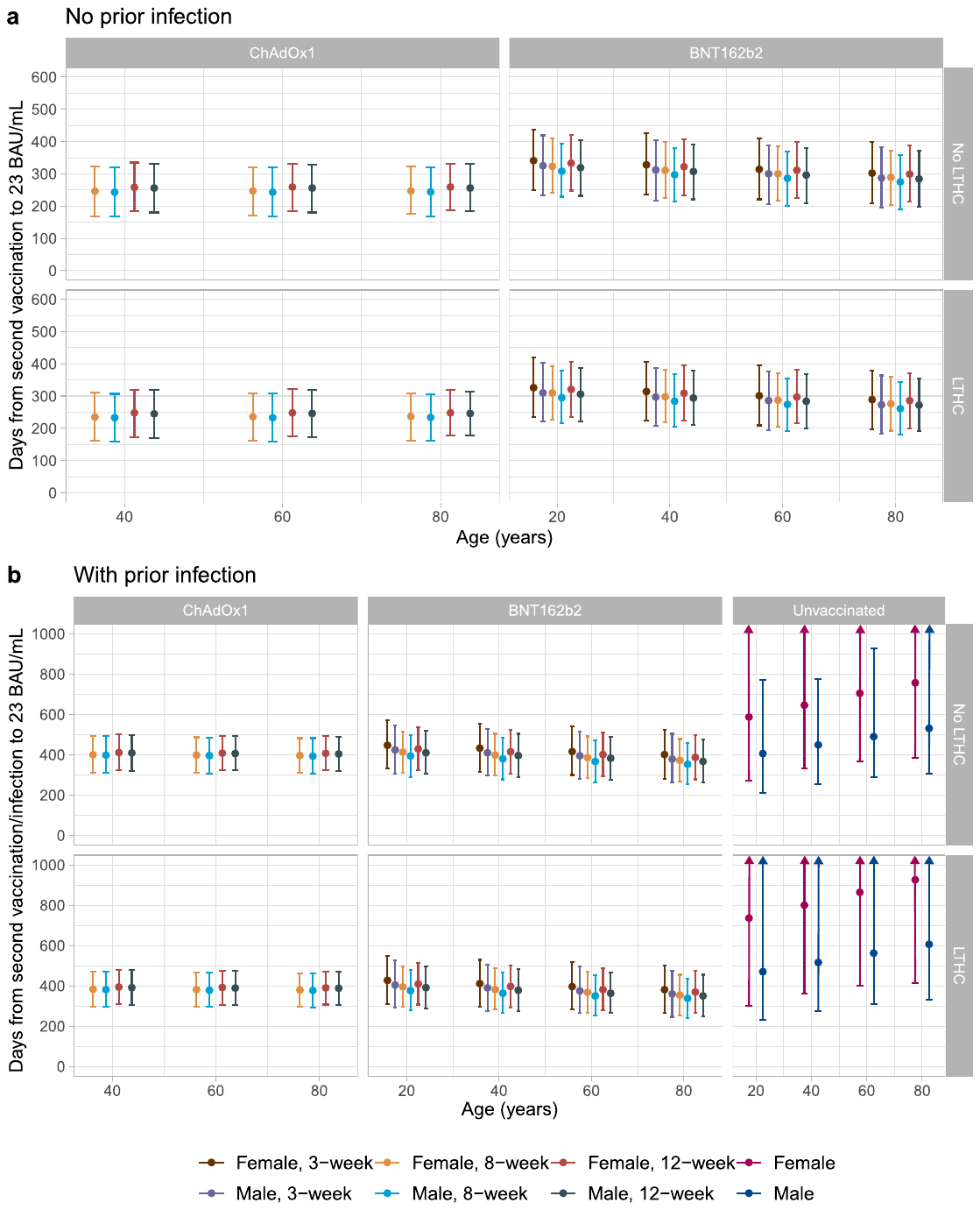

**Extended Data Fig. 7. Posterior predicted days (95% credible interval) from the second vaccination/infection to the positivity threshold of 23 BAU/mL in those without evidence of prior infection (panel a) and with evidence of prior infection, including those who had natural infection and unvaccinated (panel b).** Estimates were separated by age, sex, dosing interval, long-term health condition (LTHC), and vaccine type for vaccinated people, and by age, sex, and LTHC for unvaccinated people. y-axis is truncated at 1000 days (panel b) for visualisation. For ChAdOx1, 20-year-old group is not plotted because the vast majority of those receiving ChAdOx1 were ≥40 years. Estimates for the unvaccinated group in panel b are based on a previously reported biphasic model assuming the rate of antibody decline slowing over time^1^, which may explain the longer point estimates vs ChAdOx1 or BNT162b2, however credible intervals also overlap.

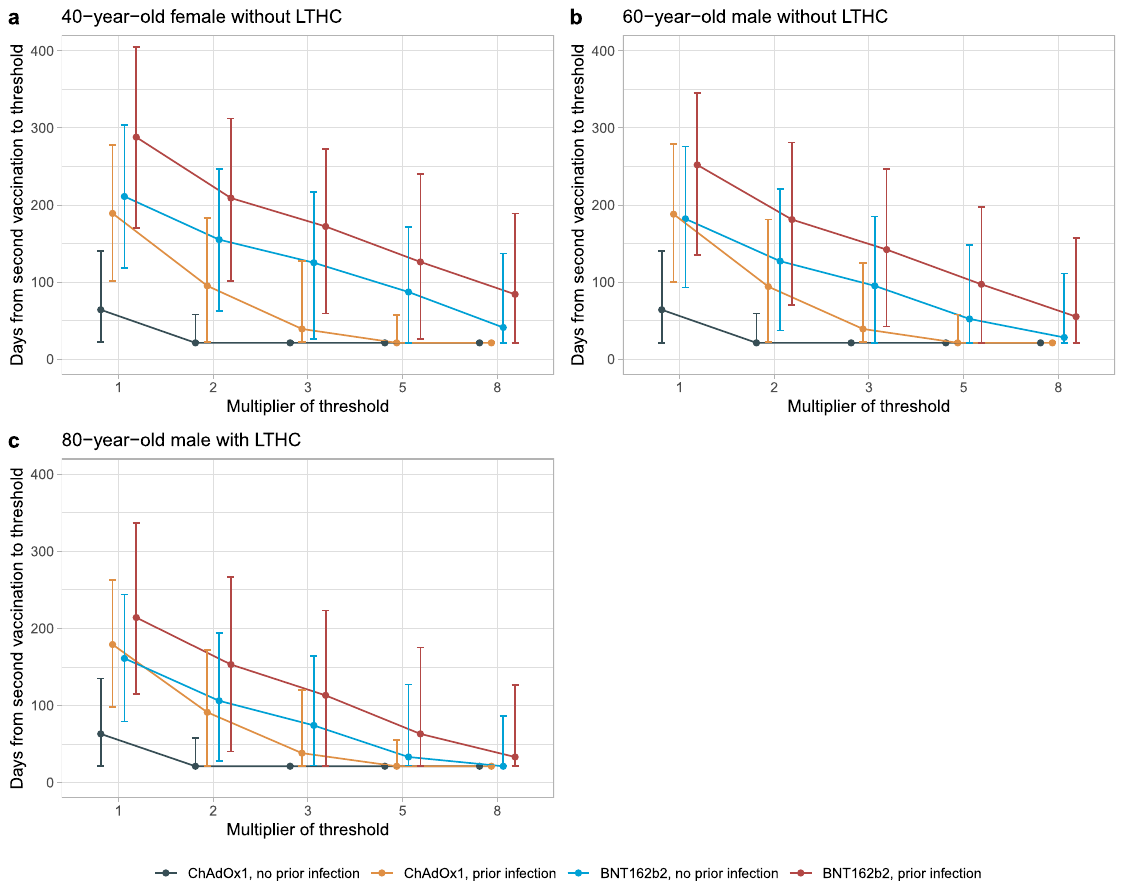

**Extended Data Fig. 8.** **Posterior predicted days from the second vaccination to the threshold level associated with 67% protection (ChAdOx1: 107 BAU/mL, BNT162b2: 94 BAU/mL) multiplied by 2, 3, 5, 8, according to prior infection status and vaccine type.** **a**, in a 40-year-old female without long-term health conditions. **b**, in a 60-year-old male without long-term health conditions. **c,** in an 80-year-old male with long-term health conditions. All three panels were plotted at an 8-week dosing interval. LTHC: long-term health condition. Multipliers reflect the fact that higher antibody level may be required for protection against variants of concern.

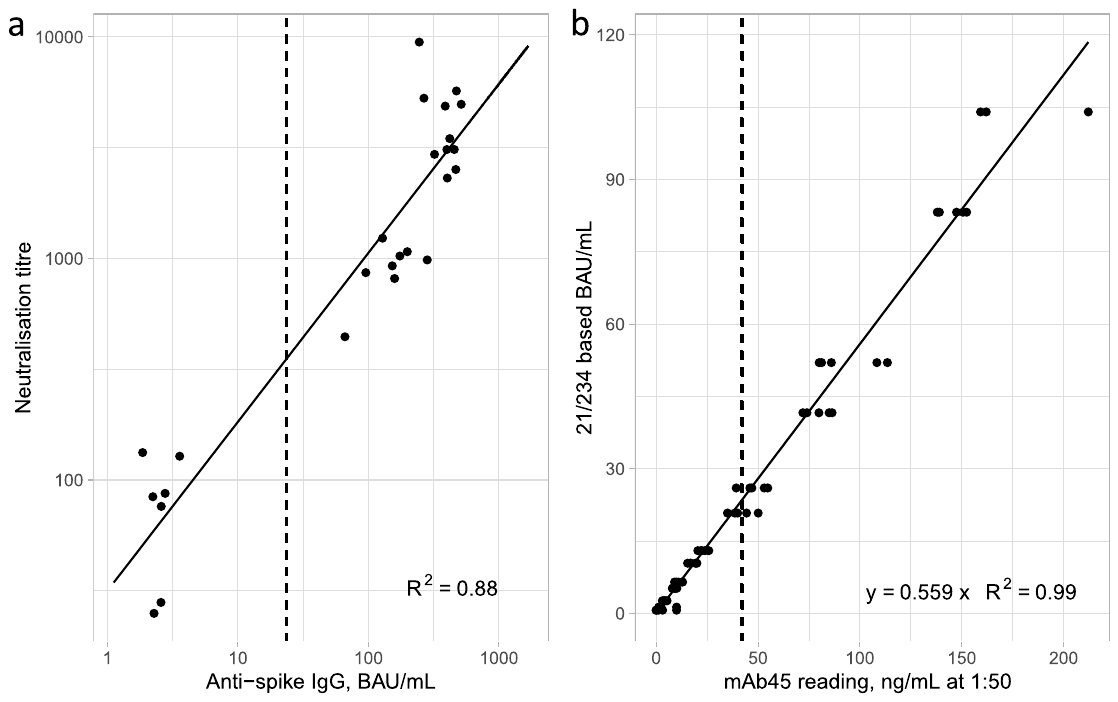

**Extended Data Fig. 9**. **a, Correlation between anti-spike IgG levels (BAU/mL) and neutralisation titres in 37 samples.** Samples were obtained from the National Institute for Biological Standards and Control (NIBSC, Potters Bar, UK) Anti-SARSCoV-2 Verification Panel for Serology Assays (NIBSC code: 20/B770). **b, Correlation between anti-spike IgG levels in mAb45 units (ng/ml) and WHO international units (BAU/mL) in 63 dilution replicates.**

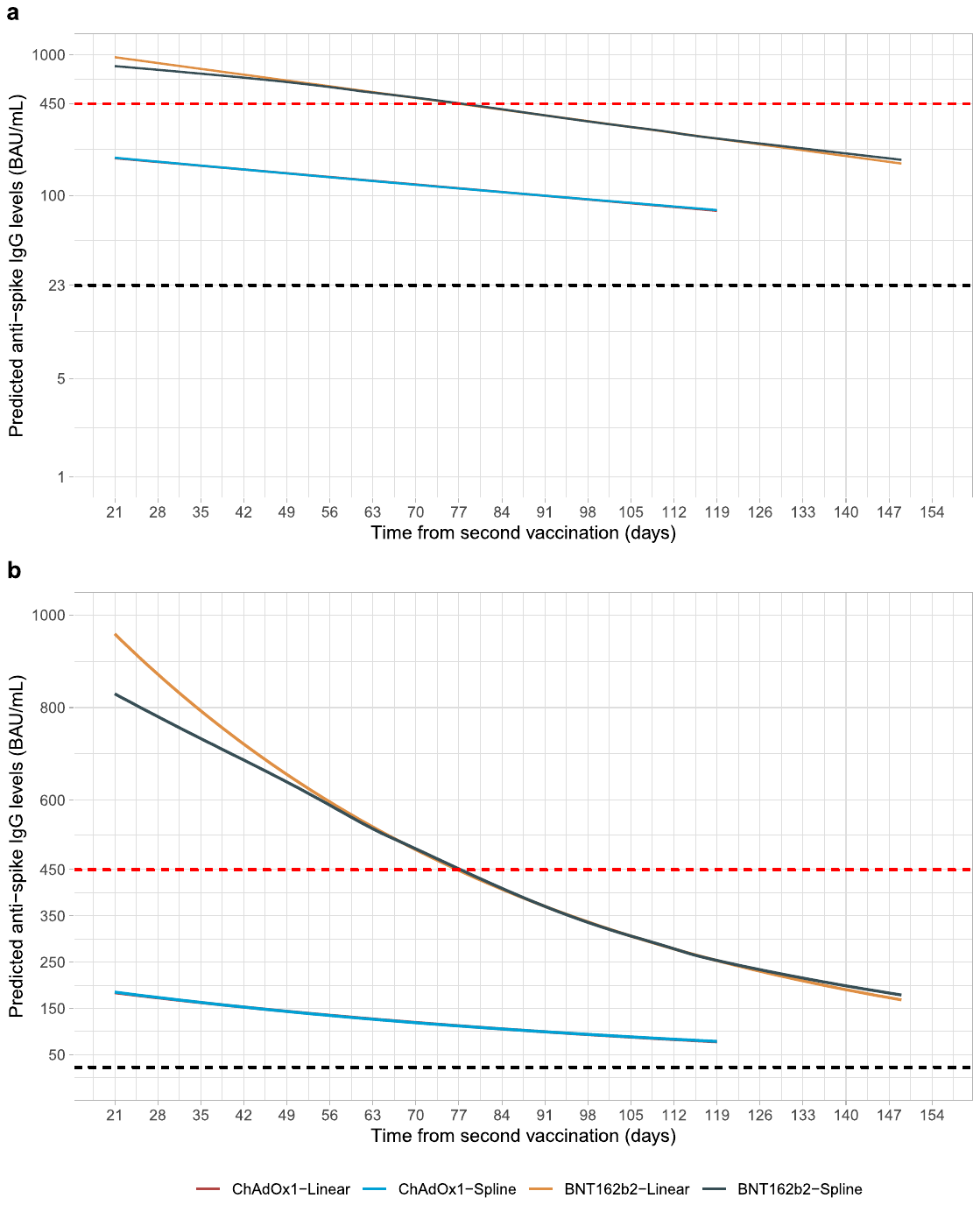

**Extended Data Fig. 10.** **Comparison of linear exponential model with spline-based model in examining non-linearity of antibody decline.** a, Plotted in log10 scale. b, Plotted in original scale. The estimated trajectory from the spline model (with 4 knots placed at 10^th^, 40^th^, 60^th^, and 90^th^ of observed time points) is similar with the linear exponential model for both ChAdOx1 and BNT162b2, indicating that there was no evidence of antibody decline flattening. Linear models provided better fit. Black dotted line shows the positivity threshold of 23 BAU/mL. Red dotted line shows the upper quantification limit of 450 BAU/mL.

### Supplementary Tables

|  | ChAdOx1 two dose no prior infection (N=121,322) | BNT162b2 two dose no prior infection (N=79,693) | ChAdOx1 two dose prior infection (N=12,066) | BNT162b2 two dose prior infection (N=9,412) | Total (N=222,493) | p value |
| --- | --- | --- | --- | --- | --- | --- |
| Percentage (%) | 54.5 | 35.8 | 5.4 | 4.2 | 100 |  |
| Dosing interval (days) | |  |  |  |  |  |
| Median | 76 | 71 | 76 | 65 | 74 |  |
| Q1, Q3 | 68, 78 | 58, 77 | 66, 78 | 56, 76 | 64, 78 |  |
| Age (years) |  |  |  |  |  | < 0.001 |
| Median | 58 | 56 | 53 | 37 | 57 |  |
| Q1, Q3 | 48, 67 | 36, 70 | 45, 62 | 29, 57 | 43, 68 |  |
| Sex |  |  |  |  |  | < 0.001 |
| Female | 64,470 (53.1%) | 44,717 (56.1%) | 6,477 (53.7%) | 5,202 (55.3%) | 120,866 (54.3%) |  |
| Male | 56,852 (46.9%) | 34,976 (43.9%) | 5,589 (46.3%) | 4,210 (44.7%) | 101,627 (45.7%) | |
| Ethnicity |  |  |  |  |  | < 0.001 |
| Non-white | 5,698 (4.7%) | 4,858 (6.1%) | 932 (7.7%) | 1,107 (11.8%) | 12,595 (5.7%) | |
| White | 115,624 (95.3%) | 74,835 (93.9%) | 11,134 (92.3%) | 8,305 (88.2%) | 209,898 (94.3%) |  |
| Household size |  |  |  |  |  | < 0.001 |
| 1 | 22,809 (18.8%) | 15,069 (18.9%) | 1,928 (16.0%) | 1,291 (13.7%) | 41,097 (18.5%) |  |
| 2 | 59,704 (49.2%) | 39,965 (50.1%) | 5,065 (42.0%) | 3,890 (41.3%) | 108,624 (48.8%) | |
| 3 | 18,176 (15.0%) | 11,691 (14.7%) | 2,167 (18.0%) | 1,859 (19.8%) | 33,893 (15.2%) |  |
| 4 | 15,215 (12.5%) | 9,143 (11.5%) | 2,075 (17.2%) | 1,593 (16.9%) | 28,026 (12.6%) | |
| 5+ | 5,418 (4.5%) | 3,825 (4.8%) | 831 (6.9%) | 779 (8.3%) | 10,853 (4.9%) |  |
| Deprivation percentile | |  |  |  |  | < 0.001 |
| Median | 64 | 62 | 60 | 56 | 62 |  |
| Q1, Q3 | 40, 83 | 38, 82 | 35, 81 | 32, 78 | 39, 82 |  |
| Report working in patient facing healthcare | |  |  |  |  | < 0.001 |
| No | 119,937 (98.9%) | 74,840 (93.9%) | 11,805 (97.8%) | 8,840 (93.9%) | 215,422 (96.8%) | |
| Yes | 1,385 (1.1%) | 4,853 (6.1%) | 261 (2.2%) | 572 (6.1%) | 7,071 (3.2%) |  |
| Report having a long-term health condition | |  |  |  |  | < 0.001 |
| No | 87,034 (71.7%) | 56,147 (70.5%) | 9,083 (75.3%) | 7,415 (78.8%) | 159,679 (71.8%) |  |
| Yes | 34,288 (28.3%) | 23,546 (29.5%) | 2,983 (24.7%) | 1,997 (21.2%) | 62,814 (28.2%) |  |

**Supplementary Table 1.** **Characteristics of participants with two vaccine doses and at least one antibody measurement from 91 days before the first vaccination through to 4^th^ October.**

| (a) |  |  |  | |  | |
| --- | --- | --- | --- | --- | --- | --- |
|  |  |  | **Without prior infection** | | **With prior infection** | |
| Time from second dose (days) | **Dosing interval (days)** | **Age (years)** | **ChAdOx1** | **BNT162b2** | **ChAdOx1** | **BNT162b2** |
| 21 | 21 | 60 | NA | 276 (236-324) | NA | 305 (212-440) |
| 21 | 56 | 60 | 159 (147-172) | 371 (340-405) | 312 (219-443) | 418 (284-614) |
| 21 | 70 | 60 | 175 (168-182) | 400 (386-415) | 325 (278-380) | 425 (355-509) |
| 21 | 84 | 60 | 169 (161-178) | 392 (367-418) | 280 (218-360) | 426 (308-588) |
| 42 | 21 | 60 | NA | 328 (284-378) | NA | 347 (243-495) |
| 42 | 56 | 60 | 132 (123-141) | 357 (324-394) | 277 (197-390) | 392 (261-587) |
| 42 | 70 | 60 | 137 (132-142) | 375 (359-391) | 294 (251-344) | 406 (342-481) |
| 42 | 84 | 60 | 143 (137-150) | 356 (330-383) | 274 (215-347) | 395 (286-547) |
| 63 | 21 | 60 | NA | 287 (249-331) | NA | 371 (260-530) |
| 63 | 56 | 60 | 109 (101-116) | 321 (295-350) | 230 (163-326) | 372 (241-572) |
| 63 | 70 | 60 | 111 (107-115) | 321 (310-333) | 254 (217-298) | 379 (318-453) |
| 63 | 84 | 60 | 121 (116-127) | 323 (304-344) | 266 (209-339) | 395 (288-542) |
| (b) |  |  |  | |  | |
|  |  |  | **Without prior infection** | | **With prior infection** | |
| Time from second dose (days) | **Dosing interval (days)** | **Age (years)** | **ChAdOx1** | **BNT162b2** | **ChAdOx1** | **BNT162b2** |
| -49 | 70 | 20 | 108 (90-130) | 295 (208-417) | 351 (317-388) | 347 (273-440) |
| 21 | 70 | 20 | 183 (154-217) | 317 (225-447) | 404 (362-450) | 420 (308-574) |
| -49 | 70 | 40 | 81 (76-87) | 247 (203-300) | 219 (208-231) | 293 (243-353) |
| 21 | 70 | 40 | 156 (148-165) | 289 (241-348) | 394 (377-413) | 410 (342-492) |
| -49 | 70 | 60 | 65 (62-68) | 255 (216-301) | 142 (135-150) | 234 (195-282) |
| 21 | 70 | 60 | 175 (168-182) | 325 (278-380) | 400 (386-415) | 425 (355-509) |
| -49 | 70 | 80 | 37 (33-42) | 112 (85-148) | 91 (83-100) | 141 (107-185) |
| 21 | 70 | 80 | 175 (160-193) | 254 (193-334) | 404 (377-432) | 422 (324-548) |

**Supplementary Table 2.** **Predicted anti-spike IgG levels (BAU/mL) with 95% confidence interval (CI) from generalised additive models. a,** Comparison of 8-, 10-, 12-, and 3-week dosing interval 21, 42, 63 days after the second dose in 60-year-olds. **b,** Comparison of 20-, 40-, 60-, and 80-year-olds 21 days after the first and second dose in those with 10-week dosing interval. The 95% confidence intervals are calculated by prediction ± 1.96*standard error of prediction. Values truncated at 450 BAU/mL counted as =450 BAU/mL.

| (a) Frequency of antibody measurements per participant | | |
| --- | --- | --- |
|  | **ChAdOx1** | **BNT162b2** |
| 1 | 37,478 | 17,863 |
| 2 | 39,268 | 16,444 |
| 3 | 20,450 | 13,839 |
| 4 | 3,442 | 6,074 |
| 5 | 1 | 832 |
| 6 | 0 | 1 |
| (b) Frequency of antibody measurements relative to the second vaccination | | |
| (days) | **ChAdOx1** | **BNT162b2** |
| 21-28 | 15,693 | 6,964 |
| 28-35 | 16,403 | 7,161 |
| 35-42 | 16,535 | 6,934 |
| 42-49 | 16,427 | 7,061 |
| 49-56 | 16,200 | 6,973 |
| 56-63 | 16,788 | 7,310 |
| 63-70 | 16,284 | 7,507 |
| 70-77 | 15,750 | 7,471 |
| 77-84 | 14,258 | 7,709 |
| 84-91 | 12,962 | 7,925 |
| 91-98 | 11,070 | 8,064 |
| 98-105 | 8,873 | 7,934 |
| 105-112 | 7,219 | 7,215 |
| 112-119 | 5,886 | 6,436 |
| 119-126 | 789 | 5,837 |
| 126-133 | 0 | 4,721 |
| 133-140 | 0 | 3,732 |
| 140-147 | 0 | 2,831 |
| 147-149 | 0 | 945 |

**Supplementary Table 3. Frequency of antibody measurements per participant and frequency of antibody measurements relative to the time of the second vaccination in the Bayesian linear mixed model**. 100,639 ChAdOx1 participants contributed 191,137 antibody measurements ≥21 days after the second dose, median (IQR) [range] 2 (1-2) [1-5] measurements per participant. 55,053 BNT162b2 participants contributed 120,728 antibody measurements ≥21 days after the second dose, median (IQR) [range] 2 (1-3) [1-6] per participant. The antibody measurements were taken a median (IQR) [range] 61 (41-83) [21-119] days after the second vaccination for ChAdOx1 and 79 (51-106) [21-149] days for BNT162b2.

|  |  | ChAdOx1 | | | BNT162b2 | | |
| --- | --- | --- | --- | --- | --- | --- | --- |
|  |  | Posterior mean | 95% Crl | | Posterior mean | 95% Crl | |
| Unadjusted baseline | Peak level (Intercept) | 184 | 183 | 185 | 959 | 944 | 974 |
|  | IgG half-life (slope) | 79 | 78 | 80 | 51 | 50 | 52 |
| Adjusted baseline | Peak level (Intercept) | 160 | 155 | 162 | 976 | 903 | 1011 |
|  | IgG half-life (slope) | 81 | 79 | 83 | 52 | 50 | 53 |
| Age | Peak level: 60 years |  |  |  |  |  |  |
|  | IgG half-life: 60 years |  |  |  |  |  |  |
|  | Change in peak level: per 10-years older | **3** | **2** | **4** | **-76** | **-84** | **-68** |
|  | Change in half-life: per 10-years older | **-1** | **-1** | **0** | 0 | 0 | 0 |
| Sex | Peak level: Female |  |  |  |  |  |  |
|  | IgG half-life: Female |  |  |  |  |  |  |
|  | Change in peak level: Male | **-5** | **-7** | **-3** | **-140** | **-164** | **-117** |
|  | Change in half-life: Male | 0 | -1 | 2 | -1 | -2 | 1 |
| Ethnicity | Peak level: White |  |  |  |  |  |  |
|  | IgG half-life: White |  |  |  |  |  |  |
|  | Change in peak level: Non-white | **41** | **36** | **47** | **141** | **78** | **208** |
|  | Change in half-life: Non-white | **-8** | **-11** | **-5** | 0 | -3 | 2 |
| Report having a long-term health condition | Peak level: No |  |  |  |  |  |  |
|  | IgG half-life: No |  |  |  |  |  |  |
|  | Change in peak level: Yes | **-5** | **-7** | **-3** | **-79** | **-104** | **-55** |
|  | Change in half-life: Yes | **-2** | **-4** | **-1** | **-1** | **-2** | **0** |
| Report working in patient-facing healthcare | Peak level: No |  |  |  |  |  |  |
|  | IgG half-life: No |  |  |  |  |  |  |
|  | Change in peak level: Yes | **10** | **2** | **19** | **287** | **221** | **358** |
|  | Change in half-life: Yes | 0 | -6 | 6 | **-5** | **-6** | **-3** |
| Deprivation | Peak level: 60 (median) |  |  |  |  |  |  |
|  | IgG half-life: 60 (median) |  |  |  |  |  |  |
|  | Change in peak level: per 10 percentile higher | **-1** | **-1** | **-1** | **5** | **1** | **10** |
|  | Change in half-life: per 10 percentile higher | **1** | **0** | **1** | 0 | 0 | 0 |
| 3-week interval | Peak level: No |  |  |  |  |  |  |
|  | IgG half-life: No |  |  |  |  |  |  |
|  | Change in ‘peak level’: Yes (vs 8 weeks) * |  |  |  | **-163** | **-227** | **-95** |
|  | Change in half-life: Yes (vs 8 weeks) |  |  |  | **6** | **2** | **10** |
| Dosing interval | Peak level: 8 weeks (median) |  |  |  |  |  |  |
|  | IgG half-life: 8 weeks (median) |  |  |  |  |  |  |
|  | Change in peak level: per 1 week longer | **6** | **5** | **7** | **12** | **1** | **23** |
|  | Change in half-life: per 1 week longer | 0 | -1 | 0 | 0 | 0 | 1 |
| Prior infection | Peak level: No |  |  |  |  |  |  |
|  | IgG half-life: No |  |  |  |  |  |  |
|  | Change in peak level: Yes | **219** | **210** | **227** | **312** | **248** | **381** |
|  | Change in half-life: Yes | **13** | **9** | **17** | **11** | **8** | **15** |

**Supplementary Table 4.** **Posterior mean and 95% credible intervals for anti-spike IgG peak level (intercept) (BAU/mL) and half-life (slope) (days) in the unadjusted models and multivariable models in 100,639 participants with two ChAdOx1 doses and 55,053 participants with two BNT162b2 doses**. The reference categories in the multivariable model are: 60-year-old, female, white ethnicity, not reporting a long-term health condition, not a healthcare worker, deprivation percentile=60, 8-week dosing interval, and no prior infection. Bold numbers indicate an effect not compatible with chance (95% credible interval excludes no effect). *The predicted peak level in 3-week vs 8-week is extrapolated and for modelling purpose only, it should not be interpreted as an actual peak.

| Days from first vaccination | ChAdOx1 | BNT162b2 |
| --- | --- | --- |
| 35-49 | 1,364 | 1,875 |
| 49-63 | 4,766 | 4,327 |
| 63-77 | 9,681 | 5,027 |
| 77-91 | 14,081 | 5,830 |
| 91-105 | 18,268 | 7,318 |
| 105-119 | 22,644 | 9,288 |
| 119-133 | 26,749 | 10,725 |
| 133-147 | 27,934 | 11,090 |
| 147-161 | 28,251 | 12,327 |
| 161-175 | 27,636 | 13,392 |
| 175-189 | 26,770 | 14,111 |
| 189-203 | 21,210 | 14,195 |
| 203-217 | 14,786 | 13,397 |
| 217-231 | 10,285 | 11,202 |
| 231-245 | 4,640 | 7,638 |
| 245-259 | 993 | 3,966 |
| 259-273 | 65 | 1,673 |
| 273-287 | 1 | 647 |
| 287-298 | 0 | 103 |

**Supplementary Table 5. Frequency of visits relative to the time of the first vaccination in the generalised additive models estimating correlates of protection**. Study visits were included from 17^th^ May 2021 to 4^th^ October 2021, i.e. while the Delta variant accounted for nearly all cases. The median (IQR) [range] was 149 (115-182) [36-273] days for ChAdOx1 and 167 (123-204) [35-298] days for BNT162b2. Note that UK started its booster programme from 16^th^ September 2021, so a small number of visits occurred in the first two weeks after initiating the booster campaign.

| a) |  | ChAdOx1 | | | BNT162b2 | | |
| --- | --- | --- | --- | --- | --- | --- | --- |
|  |  | **Total** | **Non-responder** | **%** | **Total** | **Non-responder** | **%** |
| First dose | At least one antibody measurement >=21 days after first dose and before second dose | 57,650 | 4,280 | 7.4 | 30,843 | 1,374 | 4.5 |
|  | Two or more antibody measurements, including at least one antibody measurement >=21 days after first dose and before second dose | 29,277 | 1,706 | 5.8 | 15,525 | 546 | 3.5 |
| Second dose | At least one antibody measurement >=21 days after second dose | 100,776 | 1,010 | 1 | 57,313 | 312 | 0.5 |
|  | Two or more antibody measurements, including at least one antibody measurement >=21 days after second dose | 77,947 | 496 | 0.6 | 44,311 | 146 | 0.3 |
| b) |  | **ChAdOx1** | | | **BNT162b2** | | |
| First and second dose | At least one Ab measurement >=21 days after first dose and before second dose, and after second dose | **Second dose: non-responder** | **Second dose: responder** | **Total** | **Second dose: non-responder** | **Second dose: responder** | **Total** |
|  | **First dose: non-responder** | 192 (0.5%) | 3,243 (7.2%) | 3,435 (7.7%) | 37 (0.2%) | 1,036 (5.8%) | 1,073 (6.0%) |
|  | **First dose: responder** | 99 (0.2%) | 41,212 (92.1%) | 41,311 (92.3%) | 15 (0.1%) | 16,909 (93.9%) | 16,924 (94.0%) |
|  | **Total** | 291 (0.7%) | 44,455 (99.3%) | 44,746 (100%) | 52 (0.3%) | 17,945 (99.7%) | 17,997 (100%) |
|  | Two or more antibody measurements, including at least one antibody measurement >=21 days after first dose and before second dose, and after second dose | **Second dose: non-responder** | **Second dose: responder** | **Total** | **Second dose: non-responder** | **Second dose: responder** | **Total** |
|  | **First dose: non-responder** | 59 (0.3%) | 1,092 (5.7%) | 1,151 (6.0%) | 8 (0.1%) | 364 (5.1%) | 372 (5.2%) |
|  | **First dose: responder** | 10 (0.1%) | 17,965 (93.9%) | 17,975 (94.0%) | 0 (0%) | 6,731 (94.8%) | 6,731 (94.8%) |
|  | **Total** | 69 (0.4%) | 19,057 (99.6%) | 19,126 (100%) | 8 (0.1%) | 7,095 (99.9%) | 7,103 (100%) |

**Supplementary Table 6.** **Percentages of non-responders to first or second dose of ChAdOx1 or BNT162b2 using a heuristic rule as all antibody measurements being <16 BAU/mL and having at least one antibody measurement 21 days after the first or second dose. a,** Non-responders in those who had antibody measurements after first dose or second dose. **b,** Non-responders in those who had antibodies measured after both first and second dose by responses to each dose. To examine the robustness of this definition, this rule was further applied in those having at least two antibody measurements.

|  | Responders (N=215,746) | Non-responders (N=6,747) | Total (N=222,493) | p value |
| --- | --- | --- | --- | --- |
| Dosing interval (days) | |  |  | < 0.001 |
| Median | 74 | 77 | 74 |  |
| Q1, Q3 | 64, 78 | 70, 79 | 64, 78 |  |
| Age (years) |  |  |  | < 0.001 |
| Median | 56 | 63 | 57 |  |
| Q1, Q3 | 43, 67 | 53, 72 | 43, 68 |  |
| Sex |  |  |  | < 0.001 |
| Female | 11,7859 (54.6%) | 3,007 (44.6%) | 120,866 (54.3%) |  |
| Male | 97,887 (45.4%) | 3,740 (55.4%) | 101,627 (45.7%) |  |
| Ethnicity |  |  |  | < 0.001 |
| Non-white | 12,383 (5.7%) | 212 (3.1%) | 12,595 (5.7%) |  |
| White | 203,363 (94.3%) | 6,535 (96.9%) | 209,898 (94.3%) |  |
| Household size |  |  |  | < 0.001 |
| 1 | 39,556 (18.3%) | 1,541 (22.8%) | 41,097 (18.5%) |  |
| 2 | 105,134 (48.7%) | 3,490 (51.7%) | 108,624 (48.8%) |  |
| 3 | 33,046 (15.3%) | 847 (12.6%) | 33,893 (15.2%) |  |
| 4 | 27,405 (12.7%) | 621 (9.2%) | 28,026 (12.6%) |  |
| 5+ | 10,605 (4.9%) | 248 (3.7%) | 10,853 (4.9%) |  |
| Deprivation percentile | |  |  | < 0.001 |
| Median | 63 | 60 | 62 |  |
| Q1, Q3 | 39, 82 | 36, 81 | 39, 82 |  |
| Report working in patient facing healthcare | |  |  | < 0.001 |
| No | 208,765 (96.8%) | 6,657 (98.7%) | 215,422 (96.8%) |  |
| Yes | 6,981 (3.2%) | 90 (1.3%) | 7,071 (3.2%) |  |
| Report having a long-term health condition | |  |  | < 0.001 |
| No | 156,009 (72.3%) | 3,670 (54.4%) | 159,679 (71.8%) |  |
| Yes | 59,737 (27.7%) | 3,077 (45.6%) | 62,814 (28.2%) |  |

**Supplementary Table 7. Comparison of the characteristics of responders with non-responders to first or second dose of ChAdOx1 or BNT162b2.** Non-responders were identified using a heuristic rule as all antibody measurements being <15 BAU/mL and having at least one antibody measurement 21 days after the first or second dose.

| Number of antibody measurements | 0 (N=179,855) | 1-2 (N=92,397) | 3-4 (N=78,565) | $\boldsymbol{\geq}$5 (N=51,531) | Total (N=402,348) |
| --- | --- | --- | --- | --- | --- |
| Dosing interval (days) |  |  |  |  |  |
| Median | 74 | 74 | 74 | 75 | 74 |
| Q1, Q3 | 62, 78 | 63, 78 | 64, 77 | 67, 78 | 63, 78 |
| Age (years) |  |  |  |  |  |
| Median | 55 | 54 | 56 | 60 | 56 |
| Q1, Q3 | 39, 69 | 41, 67 | 43, 67 | 49, 69 | 41, 68 |
| Sex |  |  |  |  |  |
| Female | 95,435 (53.1%) | 49,544 (53.6%) | 42,821 (54.5%) | 28,501 (55.3%) | 216,301 (53.8%) |
| Male | 84,420 (46.9%) | 42,853 (46.4%) | 35,744 (45.5%) | 23,030 (44.7%) | 186,047 (46.2%) |
| Ethnicity |  |  |  |  |  |
| Non-white | 13,874 (7.7%) | 5,099 (5.5%) | 4,521 (5.8%) | 2,974 (5.8%) | 26,468 (6.6%) |
| White | 165,981 (92.3%) | 87,298 (94.5%) | 74,044 (94.2%) | 48,557 (94.2%) | 375,880 (93.4%) |
| Household size |  |  |  |  |  |
| 1 | 36,725 (20.4%) | 15,928 (17.2%) | 14,239 (18.1%) | 10,930 (21.2%) | 77,822 (19.3%) |
| 2 | 82,100 (45.6%) | 43,323 (46.9%) | 38,341 (48.8%) | 26,960 (52.3%) | 190,724 (47.4%) |
| 3 | 27,952 (15.5%) | 14,580 (15.8%) | 12,301 (15.7%) | 7,012 (13.6%) | 61,845 (15.4%) |
| 4 | 22,978 (12.8%) | 12,970 (14.0%) | 10,027 (12.8%) | 5,029 (9.8%) | 51,004 (12.7%) |
| 5+ | 10,100 (5.6%) | 5,596 (6.1%) | 3,657 (4.7%) | 1,600 (3.1%) | 20,953 (5.2%) |
| Deprivation percentile |  |  |  |  |  |
| Median | 61 | 63 | 62 | 63 | 62 |
| Q1, Q3 | 37, 81 | 40, 82 | 38, 82 | 39, 83 | 38, 82 |
| Report working in patient facing healthcare | |  |  |  |  |
| No | 175,361 (97.5%) | 89,467 (96.8%) | 76,008 (96.7%) | 49,947 (96.9%) | 390,783 (97.1%) |
| Yes | 4,494 (2.5%) | 2,930 (3.2%) | 2,557 (3.3%) | 1,584 (3.1%) | 11,565 (2.9%) |
| Report having a long-term health condition | |  |  |  |  |
| No | 130,595 (72.6%) | 67,246 (72.8%) | 56,574 (72.0%) | 35,859 (69.6%) | 290,274 (72.1%) |
| Yes | 49,260 (27.4%) | 25,151 (27.2%) | 21,991 (28.0%) | 15,672 (30.4%) | 112,074 (27.9%) |

**Supplementary Table 8. Comparison of the characteristics of all participants ≥16 years in the COVID-19 Infection Survey who received two ChAdOx1 or two BNT162b2 vaccinations from 8^th^ December 2020 to 4^th^ October 2021 by number of antibody measurements.** 179,855 participants did not provide blood samples for antibody testing. 222,493 participants provided blood samples (92,397 had 1-2 measurements, 78,565 had 3-4 measurements, and 51,531 had ≥5 measurements) and were included in the analyses.

| Model term | Priors for ChAdOx1 | Priors for BNT162b2 |
| --- | --- | --- |
| Intercept | normal (7.5,1) | normal (9.5, 1) |
| Slope | normal (0, 0.1) | normal (0, 0.1) |
| Coefficient for change in intercept (age) | normal (0, 1) | normal (0, 1) |
| Coefficient for change in slope (age) | normal (0, 0.1) | normal (0, 0.1) |
| Coefficient for change in intercept (sex) | normal (0, 1) | normal (0, 1) |
| Coefficient for change in slope (sex) | normal (0, 0.1) | normal (0, 0.1) |
| Coefficient for change in intercept (ethnicity) | normal (0, 1) | normal (0, 1) |
| Coefficient for change in slope (ethnicity) | normal (0, 0.1) | normal (0, 0.1) |
| Coefficient for change in intercept (long-term health condition) | normal (0, 1) | normal (0, 1) |
| Coefficient for change in slope (long-term health condition) | normal (0, 0.1) | normal (0, 0.1) |
| Coefficient for change in intercept (healthcare worker) | normal (0, 1) | normal (0, 1) |
| Coefficient for change in slope (healthcare worker) | normal (0, 0.1) | normal (0, 0.1) |
| Coefficient for change in intercept (deprivation) | normal (0, 1) | normal (0, 1) |
| Coefficient for change in slope (deprivation) | normal (0, 0.01) | normal (0, 0.01) |
| Coefficient for change in intercept (3-week interval) | normal (0, 1) | normal (0, 1) |
| Coefficient for change in slope (3-week interval) | normal (0, 0.1) | normal (0, 0.1) |
| Coefficient for change in intercept (dosing interval) | normal (0, 1) | normal (0, 1) |
| Coefficient for change in slope (dosing interval) | normal (0, 0.1) | normal (0, 0.1) |
| Coefficient for change in intercept (prior infection) | normal (0, 1) | normal (0, 1) |
| Coefficient for change in slope (prior infection) | normal (0, 0.1) | normal (0, 0.1) |
| Random effect SD: intercept | cauchy(0, 0.5) | cauchy(0, 0.5) |
| Random effect SD: slope | cauchy(0, 0.01) | cauchy(0, 0.01) |
| Random effect intercept & slope covariance | lkj_corr_cholesky(2) | lkj_corr_cholesky(2) |

**Supplementary Table 9. Priors used in the Bayesian linear mixed interval-censored models.** Age (16-85 years) and deprivation percentile (0-100) were scaled by dividing by 10; dosing interval (42-91 days) was scaled by dividing by 7.

|  |  | Estimate | Est.Error | 95%Crl | | Rhat | Bulk_ESS | Tail_ESS |
| --- | --- | --- | --- | --- | --- | --- | --- | --- |
| ChAdOx1 | Intercept | 7.3174 | 0.0111 | 7.2951 | 7.3391 | 1.0023 | 1375 | 2882 |
|  | time | -0.0124 | 0.0002 | -0.0127 | -0.0121 | 1.0000 | 6611 | 5322 |
|  | age | 0.0254 | 0.0034 | 0.0187 | 0.0321 | 1.0024 | 1593 | 3235 |
|  | sex | -0.0466 | 0.0083 | -0.0631 | -0.0306 | 1.0015 | 1399 | 2808 |
|  | ethnicity | 0.3331 | 0.0202 | 0.2946 | 0.3737 | 1.0040 | 1501 | 2532 |
|  | lthc | -0.0450 | 0.0093 | -0.0632 | -0.0267 | 1.0049 | 1102 | 2022 |
|  | hcw | 0.0903 | 0.0374 | 0.0164 | 0.1634 | 1.0017 | 1509 | 2453 |
|  | deprivation | -0.0103 | 0.0016 | -0.0134 | -0.0072 | 1.0018 | 1550 | 2878 |
|  | dosing interval | 0.0524 | 0.0036 | 0.0454 | 0.0593 | 1.0020 | 1560 | 2920 |
|  | prior infection | 1.2453 | 0.0158 | 1.2146 | 1.2765 | 1.0038 | 1746 | 3266 |
|  | time:age | -0.0001 | 0.0000 | -0.0002 | 0.0000 | 1.0014 | 6671 | 6023 |
|  | time:sex | 0.0001 | 0.0001 | -0.0001 | 0.0003 | 1.0006 | 6612 | 6153 |
|  | time:ethnicity | -0.0013 | 0.0003 | -0.0019 | -0.0008 | 1.0004 | 5712 | 5471 |
|  | time:lthc | -0.0004 | 0.0001 | -0.0006 | -0.0001 | 1.0005 | 6501 | 5556 |
|  | time:hcw | -0.0001 | 0.0005 | -0.0010 | 0.0008 | 1.0005 | 6041 | 4866 |
|  | time:deprivation | 0.0001 | 0.0000 | 0.0001 | 0.0001 | 1.0007 | 7110 | 5923 |
|  | time:dosing interval | -0.0001 | 0.0001 | -0.0002 | 0.0000 | 1.0002 | 6541 | 5819 |
|  | time:prior infection | 0.0017 | 0.0002 | 0.0012 | 0.0021 | 1.0000 | 6010 | 5228 |
|  | sigma | 0.4757 | 0.0014 | 0.4729 | 0.4785 | 1.0131 | 400 | 677 |
|  | sd(Intercept) | 1.0101 | 0.0028 | 1.0045 | 1.0157 | 1.0100 | 1090 | 2678 |
|  | sd(time) | 0.0017 | 0.0002 | 0.0013 | 0.0022 | 1.0170 | 319 | 387 |
|  | cor(Intercept,time) | -0.2679 | 0.0159 | -0.2983 | -0.2367 | 1.0001 | 3372 | 4253 |
| BNT162b2 | Intercept | 9.9300 | 0.0259 | 9.8796 | 9.9810 | 1.0003 | 3553 | 4861 |
|  | time | -0.0194 | 0.0003 | -0.0200 | -0.0188 | 1.0004 | 6401 | 6408 |
|  | age | -0.1167 | 0.0065 | -0.1297 | -0.1040 | 1.0003 | 4746 | 6190 |
|  | sex | -0.2235 | 0.0187 | -0.2600 | -0.1868 | 1.0007 | 4743 | 6293 |
|  | ethnicity | 0.1944 | 0.0430 | 0.1106 | 0.2788 | 1.0010 | 4377 | 5850 |
|  | lthc | -0.1222 | 0.0196 | -0.1601 | -0.0843 | 1.0006 | 4213 | 5716 |
|  | hcw | 0.3717 | 0.0397 | 0.2961 | 0.4521 | 1.0019 | 4062 | 4894 |
|  | deprivation | 0.0075 | 0.0034 | 0.0007 | 0.0143 | 1.0006 | 4495 | 6470 |
|  | 3-week | -0.2638 | 0.0571 | -0.3754 | -0.1489 | 1.0002 | 3648 | 5632 |
|  | dosing interval | 0.0183 | 0.0085 | 0.0017 | 0.0351 | 1.0002 | 3599 | 5597 |
|  | prior infection | 0.4002 | 0.0377 | 0.3270 | 0.4747 | 1.0000 | 4529 | 5485 |
|  | time:age | 0.0000 | 0.0001 | -0.0001 | 0.0002 | 1.0003 | 7850 | 6895 |
|  | time:sex | -0.0002 | 0.0002 | -0.0006 | 0.0002 | 1.0003 | 8799 | 6686 |
|  | time:ethnicity | -0.0002 | 0.0005 | -0.0012 | 0.0008 | 1.0007 | 7573 | 6634 |
|  | time:lthc | -0.0005 | 0.0002 | -0.0009 | 0.0000 | 1.0009 | 8150 | 6849 |
|  | time:hcw | -0.0019 | 0.0004 | -0.0027 | -0.0011 | 1.0012 | 6397 | 6317 |
|  | time:deprivation | 0.0000 | 0.0000 | -0.0001 | 0.0001 | 1.0013 | 9273 | 6197 |
|  | time:3-week | 0.0019 | 0.0006 | 0.0007 | 0.0031 | 0.9999 | 5061 | 6104 |
|  | time:dosing interval | 0.0001 | 0.0001 | -0.0001 | 0.0003 | 1.0010 | 7100 | 6346 |
|  | time:prior infection | 0.0035 | 0.0005 | 0.0025 | 0.0044 | 0.9998 | 7001 | 6688 |
|  | sigma | 0.8319 | 0.0036 | 0.8248 | 0.8389 | 1.0027 | 1982 | 4821 |
|  | sd(Intercept) | 1.1414 | 0.0122 | 1.1175 | 1.1655 | 1.0032 | 2434 | 4901 |
|  | sd(time) | 0.0027 | 0.0002 | 0.0023 | 0.0032 | 1.0180 | 315 | 396 |
|  | cor(Intercept,time) | -0.9189 | 0.0447 | -0.9882 | -0.8234 | 1.0203 | 283 | 324 |

**Supplementary Table 10. Model coefficients and MCMC diagnostics for the multivariable Bayesian linear mixed models.**

### Supplementary Figures

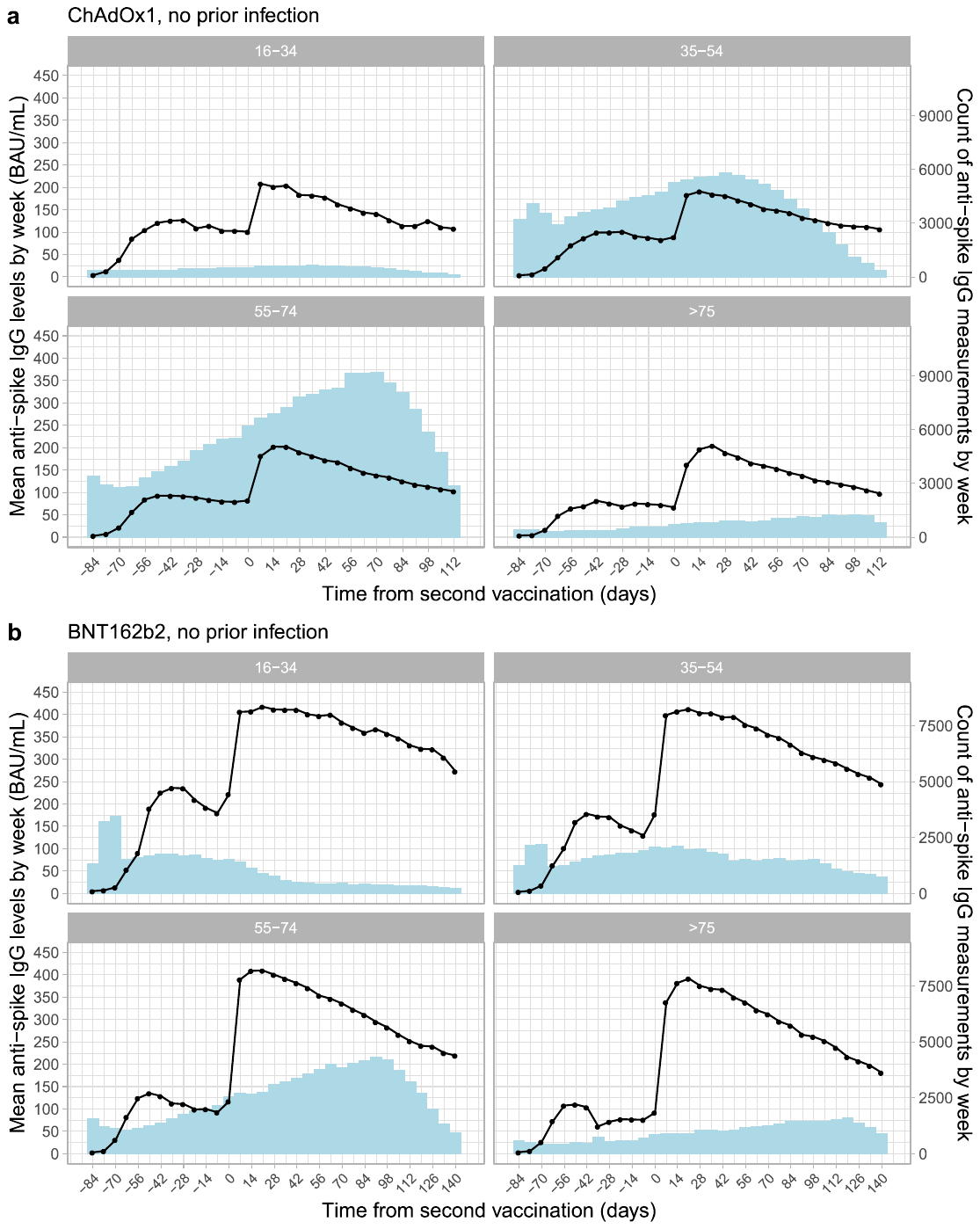

**Supplementary Fig. 1. Unadjusted mean anti-spike IgG levels (BAU/mL) and count of IgG measurements by week from second vaccination in those without prior infection.** Plots are separated by age group (16-34, 35-54, 55-74, and ≥75 years) and are not separated by dosing intervals. **a**, in those who received two ChAdOx1 vaccinations. **b**, in those who received two BNT162b2 vaccinations. Values truncated at 450 BAU/mL counted as =450 BAU/mL.

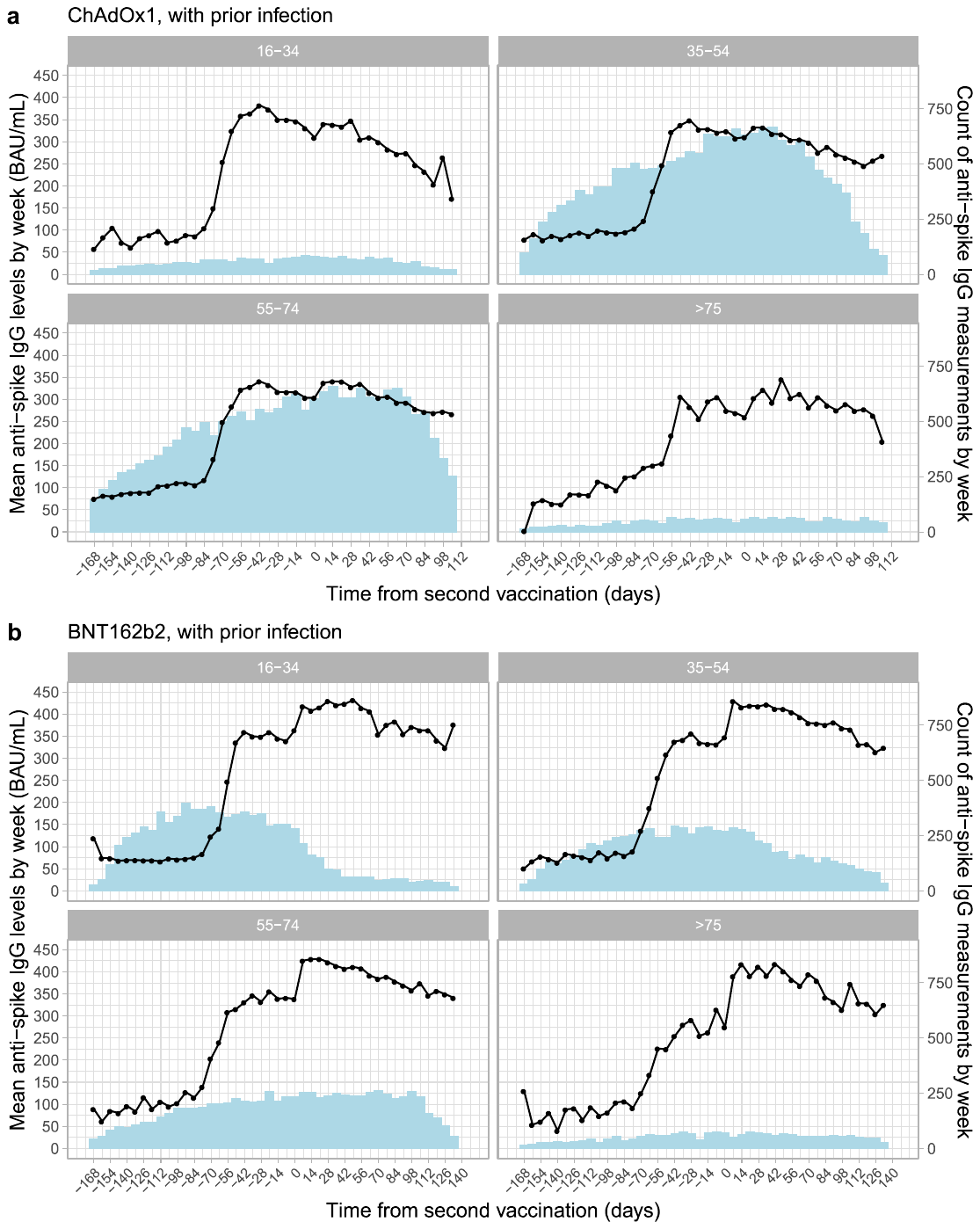

**Supplementary Fig. 2. Unadjusted mean anti-spike IgG levels (BAU/mL) and count of IgG measurements by week from second vaccination in those with prior infection.** Plots are separated by age group (16-34, 35-54, 55-74, and ≥75 years) and are not separated by dosing intervals. **a**, in those who received two ChAdOx1 vaccinations. **b**, in those who received two BNT162b2 vaccinations. Values truncated at 450 BAU/mL counted as =450 BAU/mL.

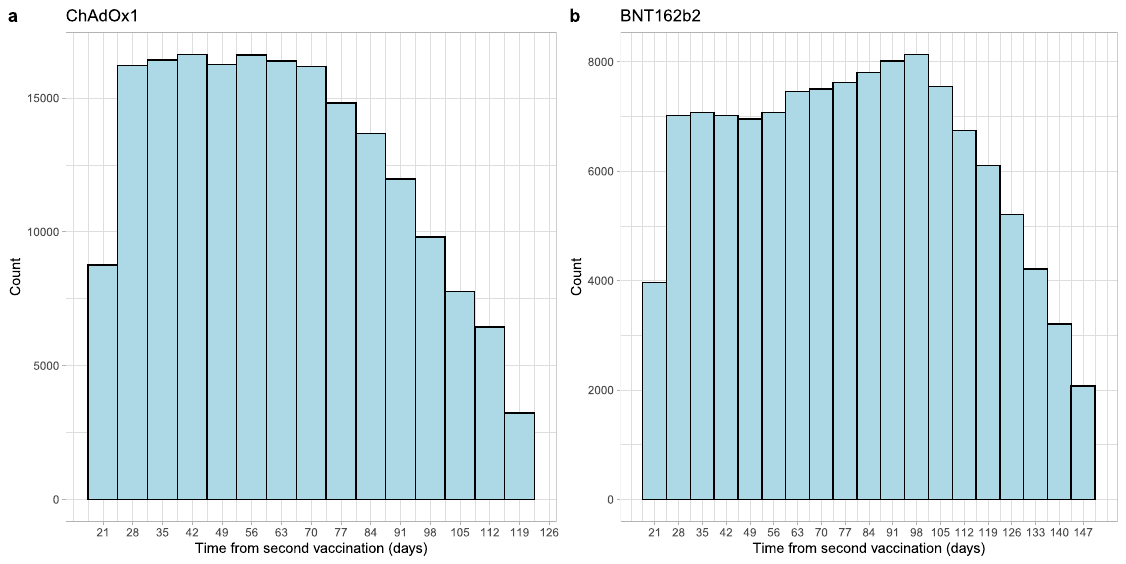

**Supplementary Fig. 3. Distribution of antibody measurements relative to the time of the second vaccination in the Bayesian linear mixed model.** The median (IQR) [range] was 61 (41-83) [21-119] days for ChAdOx1 and 79 (51-106) [21-149] days for BNT162b2.

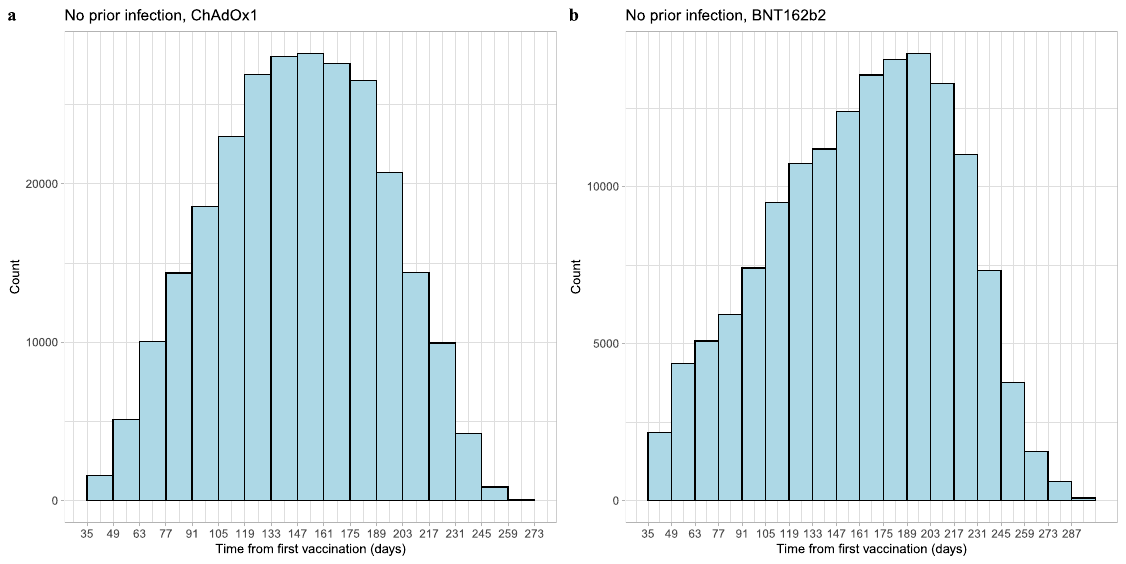

**Supplementary Fig. 4. Distribution of study visits relative to the time of the first vaccination in the generalised additive models estimating correlates of protection**. Study visits were included from 17^th^ May 2021 to 4^th^ October 2021, i.e. while the Delta variant accounted for nearly all cases. The median (IQR) [range] was 149 (115-182) [36-273] days for ChAdOx1 and 167 (123-204) [35-298] days for BNT162b2.

**References**

1. Wei, J. *et al.* Anti-spike antibody response to natural SARS-CoV-2 infection in the general population. *Nature Communications 2021 12:1* **12**, 1–12 (2021).
